# How will COVID-19 persist in the future? Simulating future dynamics of COVID-19 using an agent-based network model

**DOI:** 10.1101/2023.08.29.23294791

**Authors:** Ethan Roubenoff

## Abstract

Despite the United States Center for Disease Control (CDC)’s May 2023 expiration of the declared public health emergency pertaining to the COVID-19 pandemic (Silk 2023), approximately 3 years after the first cases of SARS-CoV-2 appeared in the United Sates, thousands of new cases persist daily. Many questions persist about the future dynamics of SARS-CoV-2’s in the United States, including: will COVID continue to circulate as a seasonal disease like influenza, and will annual vaccinations be required to prevent outbreaks? In response, we present an Agent Based Networked Simulation of COVID-19 transmission to evaluate recurrent future outbreaks of the disease, accounting for contact heterogeneity and waning vaccine-derived and natural immunity. Our model is parameterized with data collected as part of the Berkeley Interpersonal Contact Survey (BICS; Feehan and Mahmud 2021) and is used to simulate time series of confirmed cases of and deaths due to SARS-CoV-2, paying special attention to seasonal forces and waning immunity (Kronfeld-Schor et al. 2021; X. Liu et al. 2021; Nichols et al. 2021). From the BICS ABM model we simulate SARS-CoV-2 dynamics over the 10-year period beginning in 2021 with waning immunity and inclusion of annual booster doses under a variety of transmission scenarios. We find that SARS-CoV-2 outbreaks are likely to occur frequently, and that distribution of booster doses during certain times of the year—notably in the late winter/early spring—may reduce the severity of a wintertime outbreak depending on the seasonal epidemiology of the pathogen.

## 1 Introduction

Three years after the first cases of SARS-CoV-2—the pathogen responsible for the COVID-19 pandemic— appeared in the United States, many control measures put in place during the early phase of the pandemic have been eliminated (Silk 2023) in favor of a desire to return to ‘business as usual’. This includes mask mandates, shelter-in-place and work from home ordinances, physical distancing guidelines, and recommendations to isolate symptomatic cases. While vaccines for COVID-19 were a source of optimism through early 2021, it became clear over the subsequent months that waning natural and vaccine-derived immunity and the pathogen’s immunity-evading mutations rendered the vaccines less effective at ending the pandemic than initially hoped (Levin et al. 2021,Centers for Disease Control and Prevention 2021). Additionally, uptake of booster doses has lagged far behind targets (National Center for Immunization and Respiratory Diseases (NCIRD) 2022a; National Center for Immunization and Respiratory Diseases (NCIRD) 2022b). Over time, ‘pandemic fatigue’ has set in as compliance with disease-preventing behavioral mandates, especially mask usage and contact limitation, has slipped and such ordinances have been lifted (Reicher and Drury 2021). The current phase of the pandemic is substantially different from the initial phase, characterized by a highly transmissible but less severe form of the illness owing in part to many factors acting in different directions, including: highly transmissible but less severe later variants (Davies et al. 2021; Strasser et al. 2022; Yang et al. 2022), higher levels of partial or full immunity from vaccine or prior infection (Clarke 2022), higher levels of social contacts approaching pre-pandemic levels, low or absent rates of mask usage and physical distancing (Crane et al. 2021), and better treatments reducing the probability of death or severe illness after infection (National Institutes of Health 2022). While better treatments, milder variants, and prior immunity has resulted in a far lower case fatality ratio than the early days of the pandemic, avoiding the illness is still a persistent challenge for those who remain at elevated risk of severe illness and death due to COVID-19, including the elderly and people with chronic health conditions.

Understanding the various drivers of future SARS-CoV-2 outbreaks can help to plan for future interventional strategies, including vaccine distribution, school or work closure, and other non-pharmaceutical interventions. Infectious disease models can help to plan for these future outbreak scenarios by helping understand how SARS-CoV-2 will exist in our medium to long-term future. Here, we consider the effect of recurrent outbreaks, annual distribution of booster doses, and seasonal change in transmission of SARS-CoV-2. It is presently unknown if SARS-CoV-2 will demonstrate a strong seasonal pattern; however, it is hypothesized to follow the seasonal patterns of influzena and other coronaviruses (Kronfeld-Schor et al. 2021).

To answer these questions, we use a stochastic Agent-Based Network Model paramaterized with contact data from the Berkeley Interpersonal Contact Survey (BICS; Feehan and Mahmud 2021). Using BICS data allows us to consider how contact heterogeneity, household structure, and other network dynamics play into the periodicity and size of future outbreaks. Our model also includes seasonal forcing of transmission parameters, waning immunity from vaccines and prior infection, and variable-rate case importation to capture interaction with counter-seasonal populations (i.e., travel between the hemispheres experiencing opposite seasons).

Agent-Based Models are alternative to compartmental models and allow for more flexible and dynamic transmission dynamics, including network structure (Ajelli et al. 2010; Bansal, Read, et al. 2010). Roubenoff, Feehan, and Mahmud 2023 utilized a compartmental model for analyzing SARS-CoV-2 transmission. While these types of models are heavily utilized for analyzing disease transmission, one particular limitation of these models with contact data is their requirement for relatively few and well-defined categorizations. ABMs are more flexible and contrast with compartmental models by keeping track of the disease status for each individual in the simulation, rather than the tally of individuals in a particular compartment. In place of differential equations describing flows between compartments, ABMs use highly explicit (either deterministic or stochastic) rules governing interaction (Bonabeau 2002; Bruch and Atwell 2015; J. D. Baker et al. 2013; He, Ionides, and King 2010). Interactions are governed by some aspect of the simulated population contained within an objective function, such that interactions between nodes of certain values are more likely that others. As a group, ABMs are free of many of the analytic requirements of compartmental models—especially the need for of explicit transition properties between states, only an objective function for optimization— earning them the descriptor ‘plug and play’ (He, Ionides, and King 2010). Importantly for our purposes, ABMs allow for flexibility in how the population mixes, allowing for contact inequality between simulated agents through either a spatial or network component. Network models are a particular type of agent-based model that assume an explicit network structure for disease-transmitting contacts. In network models, instead of a homogenous or even matrix-structured contact pattern employed by compartmental models, disease transmission is simulated as occurring over a graph representing network connections (Bansal, Bryan T Grenfell, and Meyers 2007; Danon et al. 2011; Matt J Keeling and Eames 2005).

A plethora of agent-based and network simulations of the COVID-19 pandemic have been published and tools released. The flexible yet highly specific ways that ABMs can be used to model social interactions is ideal for testing network and behavioral interventions for COVID-19. These models include a wide set of techniques, including models that explicitly account for how individuals navigate geographic space (Cuevas 2020) and social networks (Hunter and J. D. Kelleher 2021). Agent-based models have used to estimate parameters for the COVID-19 outbreak in France (Hoertel et al. 2020), Ireland (Hunter and J. D. Kelleher 2021), and Colombia (Gomez et al. 2021). Models can utilize existing contact data, such as Moghadas et al. 2021 and Sah et al. 2021, who use data from the POLYMOD survey (Mossong et al. 2008) as inputs to an ABM to evaluate willingness to vaccinate. Holmdahl et al. 2020 test a series of behavioral interventions in nursing homes using a two-cohort ABM (patients and caregivers), finding that testing frequency and isolation are the most effective ways to limit the spread of disease. We draw on a number of general ABMs developed for COVID-19, including Covasim (Kerr et al. 2021), an aspatial model that combines Erdos-Reyni Poisson random networks with SynthPops networks that are generated from empirical contact data, and OpenABM (Hinch et al. 2021), that simulates social network interaction through stochastic network simulation at the household, occupational, and random connectivity additively.

Although it remains to be seen, research has suggested that SARS-CoV-2 may exhibit seasonality similarly to influenza and other coronaviruses, which exhibit higher incidence in the colder months (Nichols et al. 2021). Indeed, in the United States, the highest number of cases were observed in winters 2020-2021 and 2021-2022. The periodicity and severity of future SARS-CoV-2 outbreaks is currently unknown, largely since the rate of mutations and long-term vaccine-derived and natural immunity is unknown, but many mechansims are theorized (Kronfeld-Schor et al. 2021). Seasonal forcing of respiratory diseases involves a consideration of multiple temporal factors relevant to modeling the transmission of SARS-CoV-2 including seasonal changes in host behavior and immune function (Altizer et al. 2006; Grassly and Fraser 2006). Although modeling studies suggest that climate may mediate the timing and peak incidence of SARS-CoV-2 outbreaks, susceptible supply driven by population immunity is the primary driver of such dynamics (R. E. Baker et al. 2020). Many childhood diseases, namely measles, exhibit seasonal cycles driven by the birth rate and increased contact between non-immune children during the school year (Metcalf et al. 2009). In some situations, it can be assumed that the pathogen is circulating at a very low level locally in between outbreaks; in others, like influenza, seasonal human-animal interactions and case importation between populations in opposite hemispheres experiencing counter-cyclical temperature-forced outbreaks may drive timing (Lofgren et al. 2007; Lowen and Steel 2014).

We use a stochastic agent-based network simulation of SARS-CoV-2 transmission parameterized with data from the Berkeley Interpersonal Contact Survey (Feehan and Mahmud 2021) and include seasonality, annual vaccination, waning immunity, and demography. To our knowledge, this represents the first agent-based model for examining COVID-19 endemic outbreak cycles and seasonality. We find that outbreaks are likely to occur regularly and that annually-distributed booster doses can be an effective tool to eliminate regular outbreaks. Depending on seasonal epidemiology of the pathogen, booster doses are most effective when distributed at certain times of year; in the absence of seasonality, booster doses are most effective when distributed in the first half of the year, but in a seasonally-forced transmission scenario distributing vaccines in early fall is more successful at eliminating major annual outbreaks.

## 2 Methods

### 2.1 Data

Like Roubenoff, Feehan, and Mahmud 2023, we also utilize here contact survey data collected by Feehan and Mahmud 2021 as part of the ongoing Berkeley Interdisciplinary Contact Survey (BICS), which captures disease-transmitting behavior during the COVID-19 pandemic. The BICS survey, collected in several waves beginning in March 2020, is an online survey aimed at capturing the frequency and nature of respondents’ face-to-face contact over a 24-hour period. Respondents to the BICS survey are recruited through a quota sample using an online survey panel provider, Lucid. Respondents are asked to report the total number of close, face-to-face contacts they had over the previous 24 hours, and to elaborate on three such contacts in detail. Respondents are also asked to report information regarding their demographic information, household structure, and other questions regarding their behavior. We utilize responses from Wave 6 (n = 5418, 12 May - 25 May 2021) of the BICS national (U.S.) survey to capture post vaccination contact patterns.

### 2.2 Model: BICS ABM

Using Hunter, Mac Namee, and J. D. Kelleher 2017’s taxonomy for categorizing Agent Based Models, the simulation model used here is disease-specific to COVID-19 and society-specific to the behaviours captured from respondents in the BICS sample frame. Behavior is modeled on networks and is without transportation and without environment. The BICS ABM simulation population is constructed of individuals (also referred to as agents, nodes, or vertices) within households. We simulate interaction and disease spread among a population of 1000 households (approx. 3200 individuals) representative of the U.S. according to the procedure described below and in the model supplement. Each agent in the simulation directly corresponds to a respondent in the BICS or POLYMOD surveys sampled with survey weights to match the distribution of age and sex of the US population, and the agents’ demographic and behavioral data is derived from the corresponding survey respondent. The simulation includes three types of social contacts: household contacts with household members, school contacts for children below the age of 18, and non-household ‘random’ contacts. As employment data are not available for this wave of the survey, ‘random’ contacts are designed to include employment contacts for adults. Household contacts and school contacts are drawn randomly at the start of the simulation according to the procedure described below and are maintained throughout the simulation; random draws of graphs representing random non-household contacts are taken during each daytime time step. In this way, the total network is *dynamic* as it changes throughout the course of the simulation.

The graph of household contacts is drawn according to the procedure described in the model supplement, which is similar to the SynthPops procedure utilized in COVASIM (Kerr et al. 2021; Mistry et al. 2021). Briefly, a supplied number of households are created with the following two-step procedure. First, BICS survey respondents are repeatedly sampled with replacement and adjustment for survey weights to be heads of household. Heads of household are chosen to match the age- and sex-distribution of adults in the United States using 2021 American Community Survey estimates (US Census Bureau 2022). Then, households are filled by sampling BICS respondents (again, with replacement and adjustment for survey weights) who match the household head’s reported household members’ age and gender, until each household is the proper size. Respondents under the age of 18 were not ascertained in the BICS survey; instead, they are sampled uniformly from the POLYMOD UK survey (Mossong et al. 2008). Throughout the simulation, each node’s behavior is derived from the corresponding BICS survey respondent’s responses; nodes derived from POLYMOD respondents are derived from the corresponding fields in the POLYMOD survey1.

The model progresses in hourly steps through simulation time. During morning and evening hours (6pm-8am), agents have contact with all members of their household. During daytime hours, random connections occur between members of the simulation population governed by the degree (number of non-household contacts) supplied by each survey respondent. A daily contact network is drawn using the Network Configuration Model, described below. Each such contact is designated to begin at a random time during the day chosen uniformly between 8am and 6pm. Each contact has a randomly chosen duration sampled according to the following probabilities: respondents to wave 6 of the BICS survey indicated that 17.1% of contacts were less than one minute, 45.2% were between 1 minute and 15 minutes, 18.7% were less than one hour, and 18.9% were more than 1 hour. Here, we choose to use the marginal distribution rather than individual-level responses due to computational limitations. During the duration of each contact, respondents are disconnected from other members of their household and reconnected after the conclusion of the random contact. If a node is clinically infectious, they may enter isolation for the duration of symptoms; if asymptomatic, they continue to mix as before. Isolation is incorporated by a parameter representing the multiplicative reduction in random daily contacts: for example, an isolation parameter of 0.1 means that a node normally with 10 daily non-household contacts with would have 1 such contact while in isolation.

Random contacts are drawn using the Network Configuration Model, which generates a random graph of contacts that preserves each node’s degree—here, the number of daily non-household contacts. The network configuration model creates random networks using only a provided degree distribution as an input. The configuration model works through a two-step procedure (Albert-ászó Barabási 2021):

1. First, assign a degree to each node in the network such that the distribution of degrees matches the desired distribution. For each node, assign a number of ‘stubs’ or half-edges equal to the degree of that node.
2. Second, randomly and uniformly join stubs to create edges until there are no stubs remaining in the network.

Without alteration, the model may produce self-edges (a node connected to itself), or multi-edges (multiple edges connecting a pair of nodes). Sampling ‘simple’ graphs that lack self- or multi-edges is a computationally intensive procedure and non-uniform in its graph-generating process. In our application, we continue to sample graphs uniformly and remove self-edges, but maintain multi-edges. Realizations of this type are likely to have total degree less than would be implied by the supplied degree distribution.

### 2.3 Transmission of SARS-CoV-2

All agents begin susceptible and vaccine roll-out begins at the beginning of simulation time. At a given time *T* 0 a supplied number of index cases are chosen randomly to be exposed to SARS-CoV-2. In addition, a vector representing the number of cases imported daily is provided as input to the simulation—in the present application, one case weekly is imported to ensure that SARS-CoV-2 is constantly circulating at a low level. At exposure, each agent is assigned a randomly drawn number of hours spent as exposed and infectious; they then proceed to either symptomatic or asymptomatic infection with a supplied probability. Baseline probability of transmission—before considering vaccine efficacy, contact duration, non-pharmaceutical inter-ventions, and asymptomatic reduction in transmisison probability—from an infected node to a susceptible node occurs with probability *β*(*t*), where *t* is the simulation’s *t*th day in the year. The value of *β*(*t*) thus represents the probabiltiy of transmission during an hour-long contact between one clinically symptomatic node and one susceptible, unvaccinated node without mask usage.

**Figure 1:**
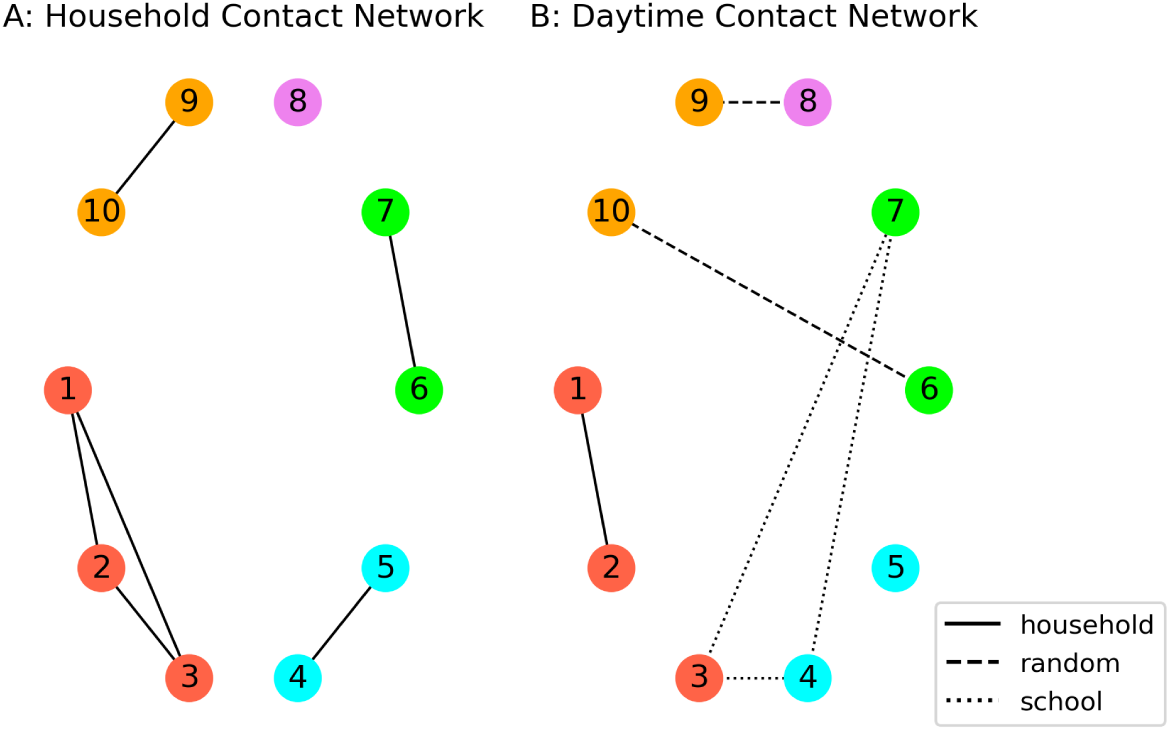
Illustration of network structures used during simulation. (A): household contact network representing evening and morning contacts, and (B): daytime contact network, consisting of school contacts and randomly drawn contacts. While school contacts are maintained throughout the simulation (with the exception of summertime school closures), random contacts are re-drawn hourly.

Various factors multiplicatively reduce the probability of transmission. First, transmission is reduced proportional to the duration of contact in fractions of an hour; a 15-minute contact is 1/4-th as likely to result in transmission as an hour-long contact. As well, transmission from an asymptomatic node to a susceptible node occurs at a reduced probability *α* relative to symptomatic nodes. The susceptible node’s vaccination status reduces the probability of transmission by the corresponding vaccine efficacy; the infectious node’s vaccination status is not assumed to affect transmission probability. As detailed further below, we allow for reinfection after a set amount of time; previous infection offers protection for the recovered node as a reduction in the transmission probability. Finally, the model includes a single Non-Pharmaceutical Intervention (NPI), designed to capture the combined disease-blocking effectiveness of masks, physical distancing, and other preventative measures. If BICS respondents corresponding to both nodes in a random contact report any mask usage, the probability of transmission is proportionally reduced by a supplied effectiveness. If NPI effectiveness is set to 0, the simulation is effectively in the absence of NPI usage. We assume that NPIs are not used among household contacts. At the conclusion of the infectious period, asymptomatic nodes all recover; symptomatic nodes die with a supplied age-based probability.

Unlike compartmental models that often hinge on parameter *R*_0_—the basic reproduction ratio, representing the average number of secondary cases caused by an index case in a fully susceptible population, estimated as the product of the transmission probability, contact rate, and duration of infectiousness—no such closed form solution for *R*_0_ in an ABM necessarily exists. Although the ABM’s ability to model contact and transmission through separate processes and objective functions allows for for increased flexibility, including time-variable and stochastic transmission probability, heterogeneous contact rates and network structure, variable duration of contact, isolation of infectious cases, and uneven NPI and vaccination usage, the parameters that govern the overall transmission dynamics are not well-defined in closed form. Instead, the *R*_0_ must be estimated from the the contact rate or incidence curve generated by the simulation itself (Hunter, Namee, and J. Kelleher 2018; Venkatramanan et al. 2018; Hunter and J. D. Kelleher 2021; Hoertel et al. 2020). We estimate *R*_0_ by observing the hourly contact rates *ĉ_h_*in the initial 7 days of the simulation, using the expression:

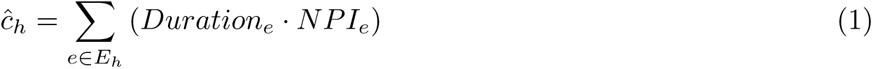

which estimates the average hourly number of edges weighted by the duration and effectiveness of non-pharmaceutical interventions, if used during each contact. Then, the average contact rate 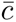 is taken to be the average of the hourly contact rates *ĉ_h_*. Finally, *R*_0_ is taken to be:

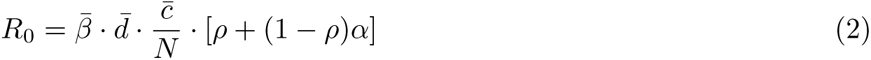

where parameter 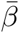 is the average transmission probability over the course of the simulation period, 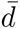 is the average duration of infectiousness, *N* is the size of the population, and the expression *ρ* +(1 *− ρ*)*α* represents the reduction in infectiousness among subclinical cases. Since *R*_0_ is not necessarily known until the conclusion of the simulation, we instead determine the overall transmission rate by varying the transmission probably *β*, then estimating *R*_0_ post-facto.

**Figure 2:**
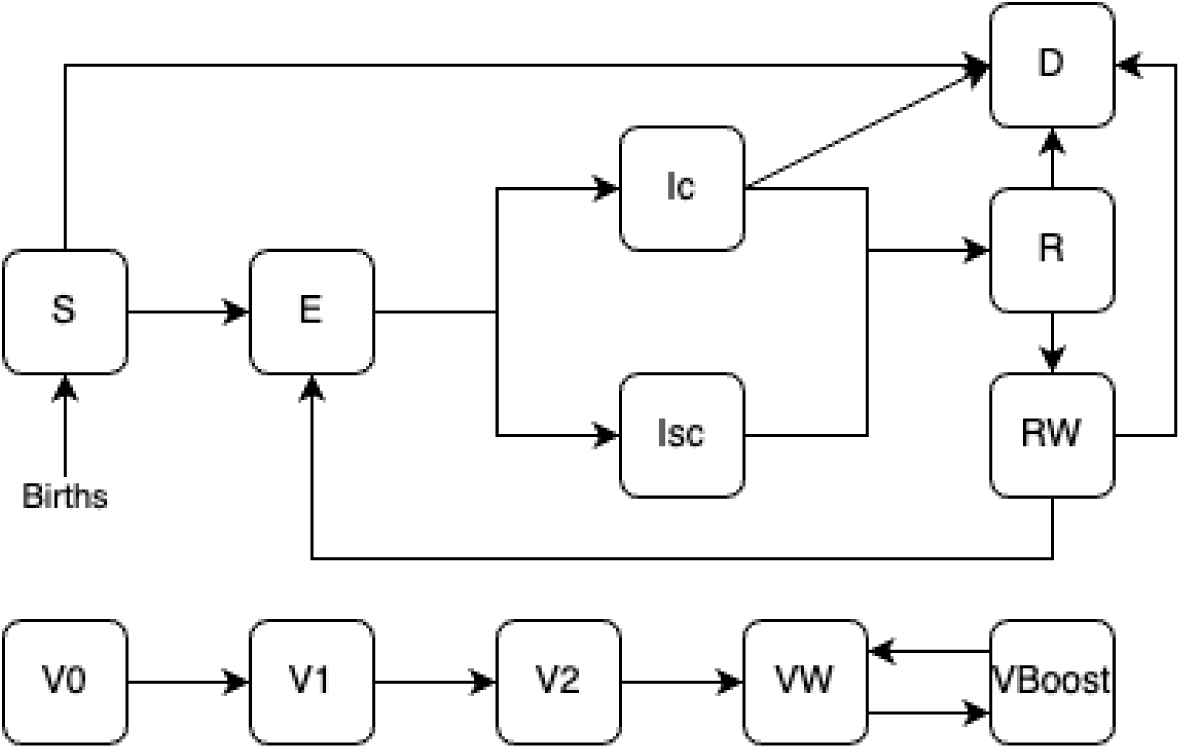
Schematic for disease states in the Agent-Based Model used in the present simulation, including disease status (top) and vaccination status (bottom). Disease states include susceptible (S), exposed/pre-infectious (E), clinically infectious (Ic), subclinically infectious (Isc), recovered (R), recovered with waned immunity (RW), and deceased (D). Vaccination states include unvaccinated (V0), first and second primary doses (V1, V2), waned immunity (VW), and boosted (VBoost).

### 2.4 Vaccine effectiveness, waned immunity, and reinfection

An important feature of the model is waning natural and vaccine-derived immunity. Vaccination occurs uniformly in the population assuming that all eligible members of the population are equally able to access the vaccine (differing from the prioritization procedure taken by Roubenoff, Feehan, and Mahmud 2023), at a baseline 70% uptake rate. A set number of vaccine doses are available daily, and are distributed at 8am each day according to optional priority rules. Nodes are eligible for a second dose of the vaccine after 25 days. After a supplied amount of time, immunity wanes, and nodes are eligible for booster doses; booster doses are made available at a set date every year and are distributed with the same rate and priority schedule as primary doses. Only nodes with waned immunity are eligible for booster doses. All vaccination statuses (1st dose, 2nd dose, waned, and booster) have fixed proportional reductions in transmission probability.

As well, we include reinfection in the model with a similar procedure. Nodes that have recovered from infection remain completely protected from infection for a fixed amount of time; after this, nodes are assigned disease status ‘Recovered-Waned’ indicating that they may be re-infected, yet have some protection against future infection. Waned immunity is assumed to be the same for clinical and subclinical infections, and the protection offered does not depend on the number of previous infections.

At present, the duration of immunity after infection and vaccination is not known, but is estimated to be approximately 6 months of near-complete immunity followed by a steady decrease over time (Centers for Disease Control and Prevention 2021). This is implemented in our model with a pair of parameters: first, the duration of complete immunity, governing the time after infection or vaccination that an individual experiences the full effect of vaccine-derived or natural immunity; second, the wanted immunity effectiveness. For simplicity, these parameters are held to be the same for both vaccine-derived and post-infectious immunity.

### 2.5 Incorporation of Seasonality

Seasonal environmental changes are known to affect infectious disease transmission in predictable, annually recurrent cycles. Although seasonality is well documented in many infectious diseases, the underlying mechanisms are frequently poorly understood or difficult to tease out from other compounding effects (Fisman 2012). For respiratory infections transmitted between humans via the airborne pathway such as SARS-CoV-2 and influenza, seasonal effects can be grouped in three broad areas: environmentally-driven changes in host behavior, such as summertime school closings or increased wintertime indoor gatherings; the pathogen’s ability to survive outside of the human host adapted to certain climatic conditions, in turn affecting fitness for transmission; and seasonal changes in the host’s immune response, possibly due to changes in temperature or sunlight exposure (Altizer et al. 2006; Grassly and Fraser 2006; Dowell 2001; Kronfeld-Schor et al. 2021; Buonomo, Chitnis, and d’Onofrio 2018; Held and Paul 2012). Additionally, seasonal migration—especially temporary domestic migration with an annual cyclical pattern—may fundamentally change the landscape of interactions and population at risk (Buckee, Tatem, and Metcalf 2017).

Incorporating seasonality into a compartmental model is typically done by adding sinusoidal temporal forcing to the transmission parameter *β* as *β*(*t*) = *β*_0_(1 + *β*_1_*cos*(2*πt*)), through a binary indicator in the case of seasonal school closings as *β*(*t*) = *β*_0_(1 +*β*_1_term(*t*)), or other time-dependent functional form (Grassly and Fraser 2006; Matt J. Keeling, Rohani, and Bryan T. Grenfell 2001). Here, the basic reproductive number *R*_0_ represents the average number of secondary cases from a single index case introduced at a random time of the year, and is defined as 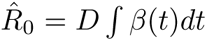 where D is the average duration of infection. Forcing functions can be easily extended to include age-structured contact (Bolker and B. Grenfell 1993), and time-series methods allow for modeling of outbreak dynamics without fitting a functional form to the transmission parameter (Metcalf et al. 2009; Finkenstádt and B. T. Grenfell 2000). Extending beyond compartmental models, seasonality can be incorporated into modeling of incidence data; Held and Paul (2012) demonstrate how seasonal incidence can be decomposed into an endemic and epidemic component with independent temporal structure.

The long-term seasonal patterns and drivers of COVID-19 are still unknown and not necessarily possible to disentangle from control efforts, especially noting non-pharmaceutical interventions like shelter-in-place ordinances and mask usage. Weaver et al. 2022 note in a review that COVID-19 may be more stable and more transmissible in cooler environments, consistent with influenza2, although both stability in high humidity and low humidity have been observed (Morris et al. 2021; Marr et al. 2019; Matson et al. 2020; Dabisch et al. 2021). SARS-CoV-2’s preference for colder, drier conditions is consistent with climatic effects observed with influenza (Lofgren et al. 2007; Lowen and Steel 2014; Shaman and Kohn 2009). Another review article by Mecenas et al. 2020 finds a similar conclusion. While climate may affect SARS-CoV-2’s transmissibility directly, the indirect effect of climate’s effect on human behaviors has been demonstrated to be a much stronger effect (Susswein, Rest, and Bansal 2023; Damette, Mathonnat, and Goutte 2021; Weaver et al. 2022). Indeed, research on contact patterns that relate to COVID-19 have are known to be a substantial driver of outbreak dynamics (Feehan and Mahmud 2021).

While many applications of ABMs to infectious disease focus on investigating the interaction-level, network, or transportation aspects of infectious disease, few have focused directly on seasonality. Arduin et al. 2017 incorporate seasonal forcing of pneumococcal infections linked to influenza infection using a fixed multiplication of the transmission probability during the flu season, similar to the school-term forcing of the transmission probability used in compartmental models by Matt J. Keeling, Rohani, and Bryan T. Grenfell 2001. Similarly, Williams et al. 2022 incorporate seasonal forcing to an ABM used to study influenza by adding a multiplicative effect to the transmission parameter related to the temporal distance of each time period from the winter solstice, which is ‘intended to account for factors that may influence transmissibility across a range of seasons due to variability in factors such as temperature, humidity, and changes in contact rates.’ In an application of ABMs to COVID-19, Krivorotko et al. 2022 use an time-series model to decompose incidence counts into a time series effect, a seasonal effect, and a noise component; the seasonal effect and the time-series effects are specified to have a temporally autocorrelative function. ABMs have also been used to study seasonality in non-human diseases (Dawson et al. 2018; Oraby et al. 2014).

We incorporate seasonality in two ways: in the transmission probability *β*(*t*) and in the number of nonhousehold contacts. We allow for seasonal forcing of the transmission probability to capture how the transmissibility of the pathogen may change with weather, modeled as:

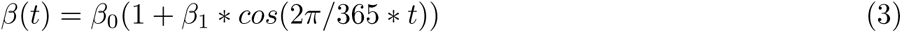

where *β*_0_ represents the average transmission probability and *β*_1_ represents the amplitude of seasonal forcing (X. Liu et al. 2021; Grassly and Fraser 2006). Second, we include school contacts between children under 18, which are derived from the POLYMOD survey (Mossong et al. 2008). School contacts are drawn once at the start of the simulation and maintained through simulation time. Schoolchildren are taken to have school contacts during the same business hours as random contacts between September 1st and June 1st annually; children who are clinically infectious with SARS-CoV-2 are kept home from school until they recover.

### 2.6 Incorporation of Demography

Optional in the model is the inclusion of basic demographic vital rates in the form of age-specific fertility and mortality data. At baseline, we draw from the CDC’s 2021 estimates (Martin, Hamilton, and Osterman 2022; Xu et al. 2022) summarized to the age categories used in the model (0-18, 18-25, then in 10 year increments through age 85). The rates used at baseline are shown in figure 12. These rates are used to randomly introduce new susceptibles into the population and randomly remove members of the population, representing deaths due to non-SARS-CoV-2 causes. Each birth represents a new, fully susceptible and unvaccinated child in the population in the household of the birthing parent; they are assumed to have the number of non-household and school contacts equal to the population average. Demography is incorporated into the model once monthly as a Bernoulli random draw for each member of the population with rate equal to the supplied rate, divided by 12 (for males, fertility rate is 0).

### 2.7 Simulation Procedure and Parameters

We identify a set of baseline transmission parameters in line with those used by Roubenoff, Feehan, and Mahmud 2023, adapted to fit the parametric needs of our Agent Based Model. For all simulations, the number of households is fixed at 1, 000, producing approximately 3, 200 individuals at the start of the simulation. Simulations are run for 10 years and are seeded with 5 index cases at time *t* = 0, intended to mimic the wintertime outbreak of early January 2021. During the course of the simulation, one case is imported weekly to ensure that all SARS-CoV-2 is constantly circulating at a low level. To account for a wide range of transmission scenarios, we consider three levels of transmissibility: low, with *β*_0_ = 0.01 (implying *R*_0_ *≈* 1.3); moderate, with *β*_0_ = 0.025 (*R*_0_ *≈* 3.4); and high, with *β*_0_ = 0.05 (*R*_0_ *≈* 6.5). Baseline simulations are without seasonal forcing of transmission or contact but with school contacts included during school-year weekdays (Monday-Friday, 9am-3pm). Seasonal forcing of both transmission and contact parameters is introduced with low (10%), moderate (25%), or extreme (50%) seasonal amplitude as described above. To introduce stochasticity into the model, each infected case in the simulation contains a randomly drawn duration of latent period of between 48 and 96 hours after transmission; this is followed by a randomly drawn infectious period of between 72 and 120 hours. These distributions are held constant across all simulations. We assume that each case has a 20% chance of being subclinical—fewer than used by Roubenoff, Feehan, and Mahmud 2023 (derived from Johansson et al. 2021), but in line with recent meta-review estimates by Buitrago-Garcia et al. 2022. That same analysis identified seven studies comparing the secondary attack rate of asymptomatic cases and symptomatic cases with an average ratio of 32%. At baseline we assume that symptomatic cases have all of their normal random contacts and do not isolate, for a ‘business as usual’ scenario; we test the effect of isolation in sensitivity analysis. However, children are assumed to always be kept at home when ill. NPI effectiveness is set to zero, equivalent to the absence of NPIs or masks, but is varied during sensitivity analysis.

Vaccine effectiveness is taken to be 80% after one, dose, 90% after two doses, and 80% after three doses, consistent with estimates published in 2021 and 2022 (Tenforde 2021,Thompson 2021; Thompson 2022) and values used by Roubenoff, Feehan, and Mahmud 2023. Unclear at the present moment is the duration of immunity after infection and vaccination and the effectiveness of waned immunity; a 2021 CDC brief (updated 2022) estimates 6 months of nearly complete immunity that diminishes over time (Centers for Disease Control and Prevention 2021). We take 6 months to be the baseline assumption but vary this duration in monthly increments from 6 months to two years in a sensitivity test. We assume a pessimistic 25% efficacy of waned natural- and vaccine-derived immunity.

All main simulations are run for 10 years in replicates of 10, and we report a number of summary values for all simulations. These include the total number of clinical and subclinical cases, deaths due to SARS-CoV-2, and the timing and size of all outbreaks after year 5. All estimates are standardized to the population size to account for populations that vary randomly in size. Outbreaks are found using the `Python` library `scipy`’s `find peaks` function on the daily sequence of clinical cases, for a minimum incidence threshold of 5% of the population infected over a 30-day window.

## 3 Results

To elucidate future outbreaks of SARS-CoV-2, we simulate outbreaks at various levels of transmissibility, and test the distribution of annual booster doses in the absence of and presence of seasonality. We find that the optimal date of booster dose distribution for reducing the number of clinical infections is different for the simulations with and without seasonality; in the absence of seasonality booster doses in the first half of the year are most effective at eliminating a large annual outbreak, but with seasonality booster doses are most effective when distributed in early fall.

Infectiousness of COVID-19 and the duration of immunity after infection and vaccination have a strong effect on the dynamics of outbreaks. In a moderate transmission scenario, where the base probability of transmission for an hour-long contact the absence of NPIs or vaccination *β*_0_ = 0.025 (corresponding to an *R*_0_ of approximately 3.2), an average of 6.16 clinical infections occur per capita over a 10-year period. Over this period, an average of 33.6 outbreaks occur, each infecting an average of 17.7% of the population.

However, when *β*_0_ is raised to 0.05 (corresponding *R*_0_ *≈* 6.5), outbreaks are fewer (an average of 16.0 over the 10-year period) but more severe, with an average outbreak size of 59.63% and about 9.75 infections per capita—nearly one per person per year. These simulations are summarized in figure 3 and trajectories are shown in figure 4. We observe this dynamic throughout many simulations: when SARS-CoV-2 is more transmissible or less mitigated, outbreaks are fewer but more severe. Mortality in the high-transmission scenario is higher, proportional to the number of clinical infections—2.63% on average compared to 1.67% in the moderate transmission scenario and 1.1% in the low-transmission scenario. These simulations suggest that the public health planning and response for future variants may differ based on their epidemiology. More transmissible variants are likely to have fewer, larger outbreaks that may overwhelm the healthcare system capacity, but show few cases outside of the season; less transmissible variants may require a year-round response, but with less severe outbreaks.

**Figure 3:**
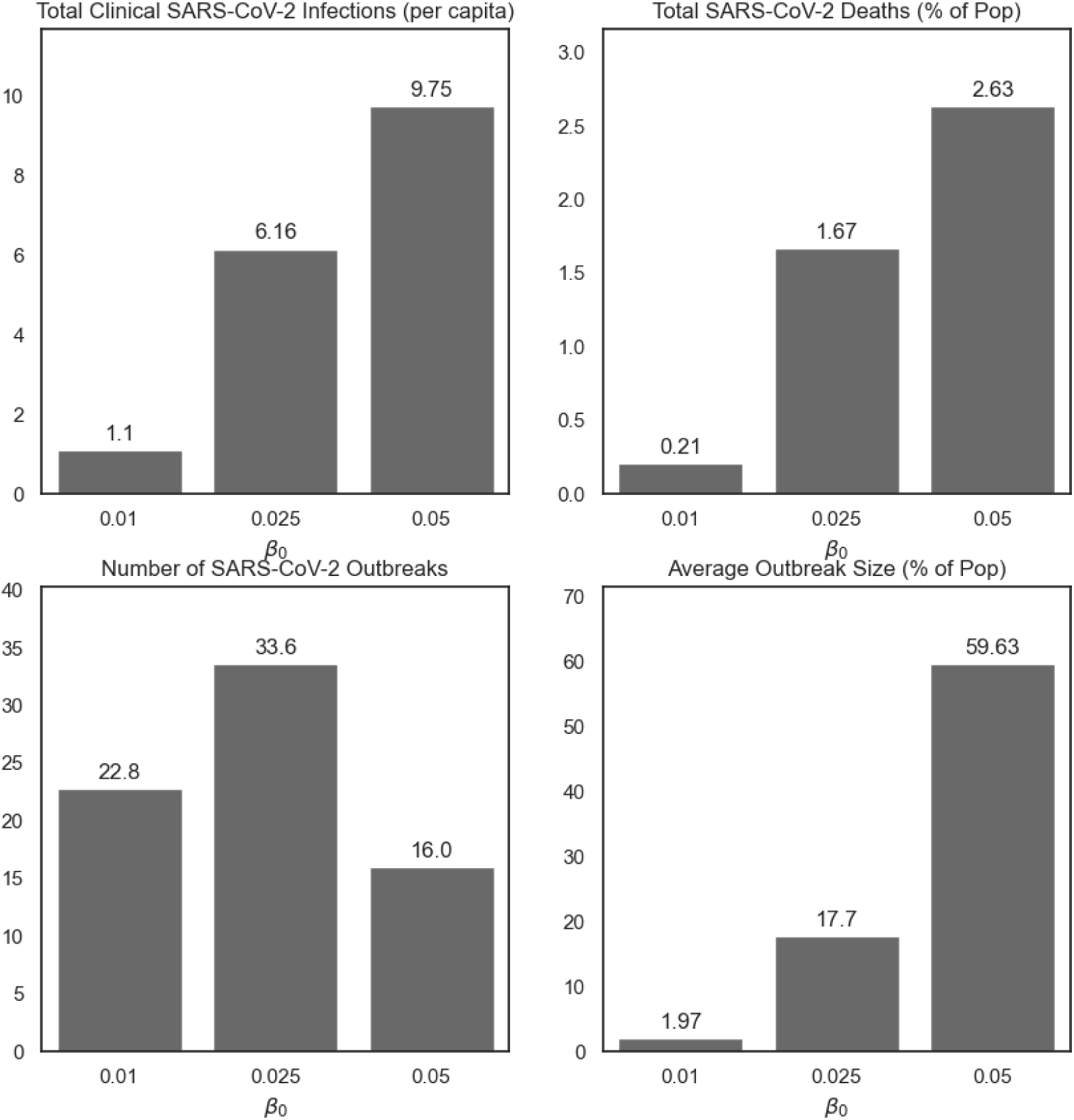
Summary of SARS-CoV-2 clinical infections, deaths, number of outbreaks, and average outbreak size for different values of *β*_0_, the average baseline transmission probability, in the absence of seasonal forcing, isolation, or vaccine distribution. Simulations are run for 10 years in replicates of 10. Approximate corresponding values of *R*_0_ are: *β*_0_ = 0.01, *R*_0_ *≈* 1.3; *β*_0_ = 0.025, *R*_0_ *≈* 3.4; *β*_0_ = 0.05, *R*_0_ *≈* 6.5.

**Figure 4:**
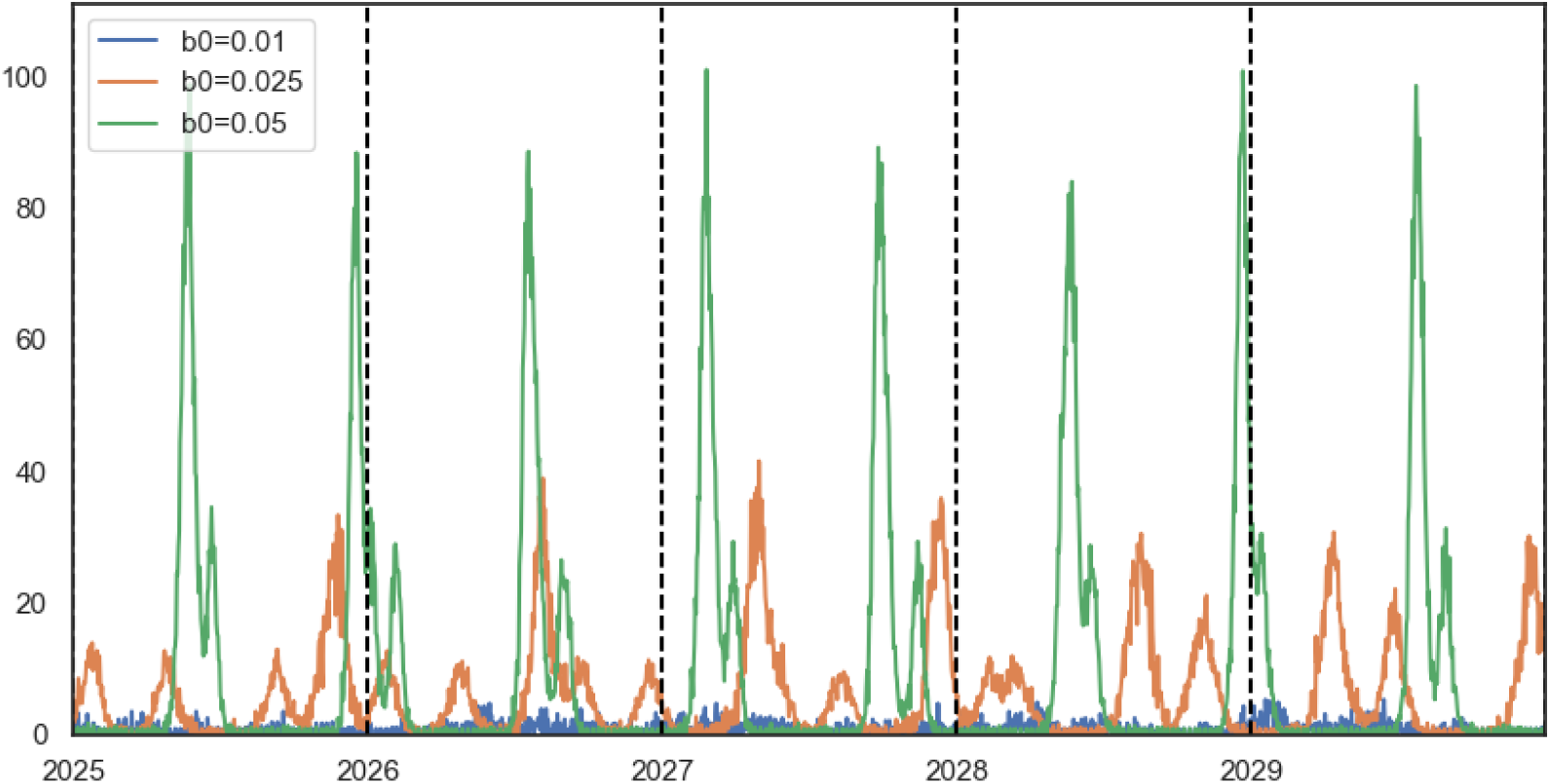
5-year trajectory of SARS-CoV-2 clinical infections for different values of *β*_0_, the average baseline transmission probability, averaged across 10 simulations each. When *β*_0_ is high, there are generally 1-2 large outbreaks per year; when lower, outbreaks are smaller and more frequent. Approximate corresponding values of *R*_0_ are: *β*_0_ = 0.01, *R*_0_ *≈* 1.3; *β*_0_ = 0.025, *R*_0_ *≈* 3.4; *β*_0_ = 0.05, *R*_0_ *≈* 6.5.

Distributing annual booster shots consistently lowers the rates of SARS-CoV-2 for all simulations, but may independently induce a seasonal pattern in outbreaks. The ability of booster doses to successfully limit SARS-CoV-2 outbreaks is dependent on the timing of their distribution and whether or not seasonal forcing of transmission is included in the model. In the absence of seasonal forcing of transmission and contact (figures 5 and 6), distributing booster doses earlier in the year is most effective at reducing the size of outbreaks: assuming a 75% update of vaccinations distributed annually between January and May, about 1.68 *−* 2.35 clinical infections occur per capita (average outbreak size ranging from 4.25% to 6.13% of the population); when distributed after July 1st, this rises to 4.34 *−* 4.76 clinical infections per capita (average outbreak size ranging from 13.21% to 15.61%). In a high-vaccine uptake scenario (90% uptake, shown in the model supplement), the overall rates of SARS-CoV-2 are lower only when vaccines are distributed in the first half of the year; when vaccines distributed in the latter half of the year, the total cases and severity of outbreaks is comparable to the regular-uptake scenario. Outbreaks are observed to generally occur around 6 months—which is also the duration of full immunity after vaccination—after the date that booster doses are made available, with the strongest seasonal patterns observed with June-September distribution inducing a strong wintertime and October - December inducing an early spring outbreak. Indeed, as observed in figure 6, the peak out the outbreak appears to be ‘shifted’ approximately 6 months after the date of booster dose distribution, the duration of of full immunity after vaccination. This phenomenon—whereby January-May boosters nearly eliminate annual outbreaks in the steady state but later-year boosters fail to do so, albeit shift the outbreak timing—is likely driven by the initial outbreak (set to occur in January of 2021 to capture the large winter outbreak in the United States), which establishes the clock for a sufficient number of individuals with waned immunity to appear for an outbreak to occur predictably after. These simulations indicate that, in the absence of seasonality, the timing of booster dose distribution may have the power to govern the timing of the primary annual outbreak.

**Figure 5:**
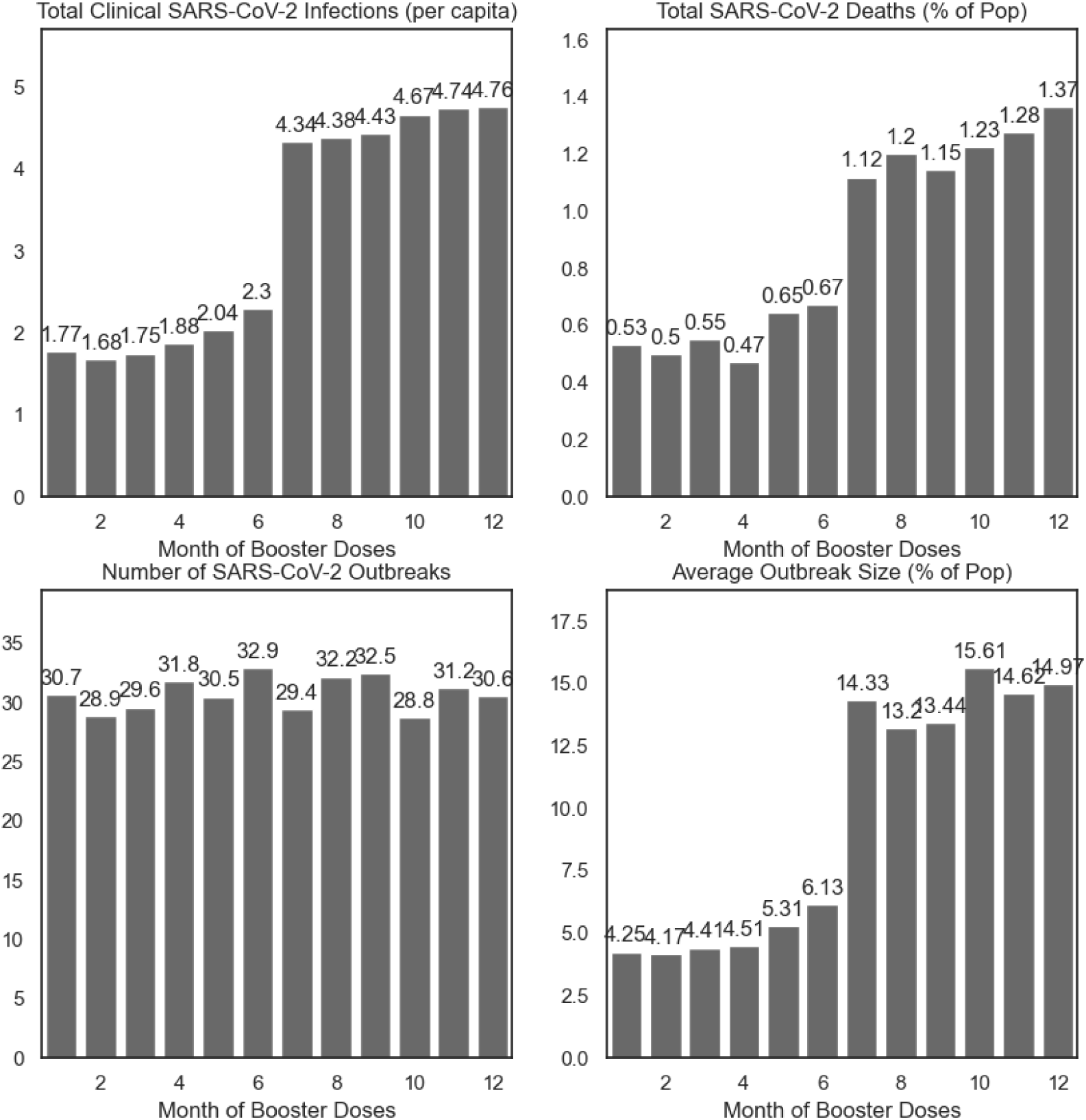
Summary of simulations by day of booster dose distribution, varied as the first of each month, in the absence of seasonal forcing of the transmission parameter *β*. Simulations are run for 10 years in replicates of 10.

**Figure 6:**
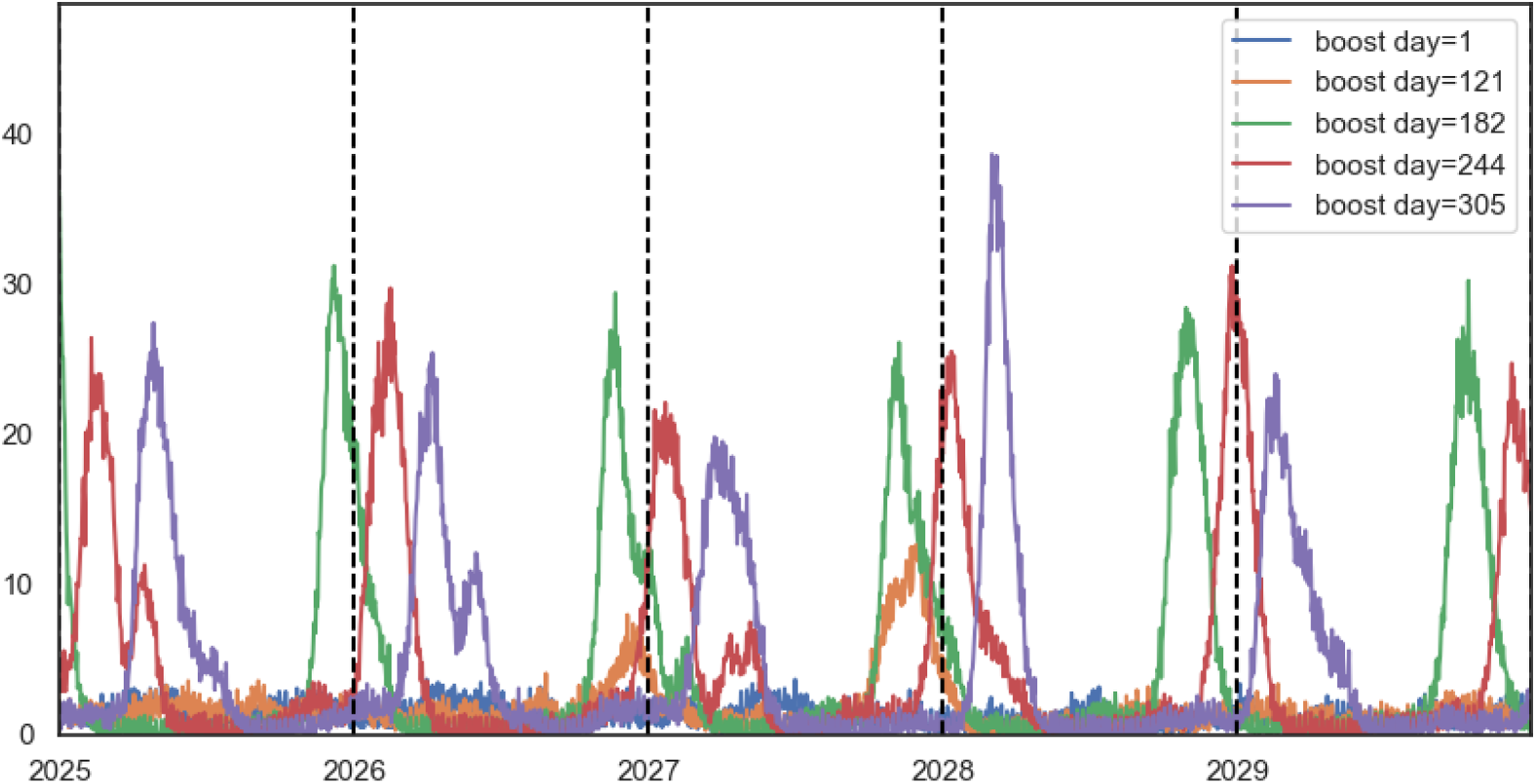
5-year trajectory of simulations by day of booster dose distribution, in the absence of seasonal forcing, for selected distribution days: Jan 1st (day 1), May 1st (day 121), July 1st (day 182), Sept 1st (day 244), and Nov 1st (day 305), averaged across 10 replications. When doses are distributed earlier in the year—Jan 1st-May 1st—the major outbreaks are largely averted, but persist when doses are distributed too late in the year.

We also tested the distribution of vaccines in the presence of seasonal forcing of the transmission parameters (*β*_1_ = 0.5), shown in figure 7 and 8. Unlike when distributing booster doses in the absence of seasonal forcing, in which the timing of booster dose distribution shifts the timing of the main outbreak, in these simulations with seasonal forcing a substantial wintertime outbreak occurs at nearly the same time every year. However, the size of this outbreak, as well as the presence of secondary outbreaks throughout the year, depends on the timing of booster doses. When boosters are distributed in the first half of the year— January 1st through May 1st—a moderate-sized fall outbreak occurs. When boosters are distributed by July 1st, this fall outbreak nearly doubles in size; a September 1st distribution day results in a less predictable situation, with multiple (2-3) smaller outbreaks throughout the year. However, distributing doses too late (November 1st) results in a large summertime outbreak, despite the relatively lowered transmission rate during the summer months. These dynamics are similar when seasonal forcing is present in the transmission parameters and the contact rate, shown in the supplementary material. Overall, simulations where vaccines are distributed between January and June resulted in 2.76 *−* 3.36 infections per capita—1-1.5 fewer than when vaccines are distributed in the highest months of July or December (4.82, 5.19 respectively). However, distributing booster doses in September or October results in fewer infections, comparable with simulations when vaccines distributed earlier in the year, and smaller outbreaks (8.69%*−*9.59% of the population infected on average per outbreak).

**Figure 7:**
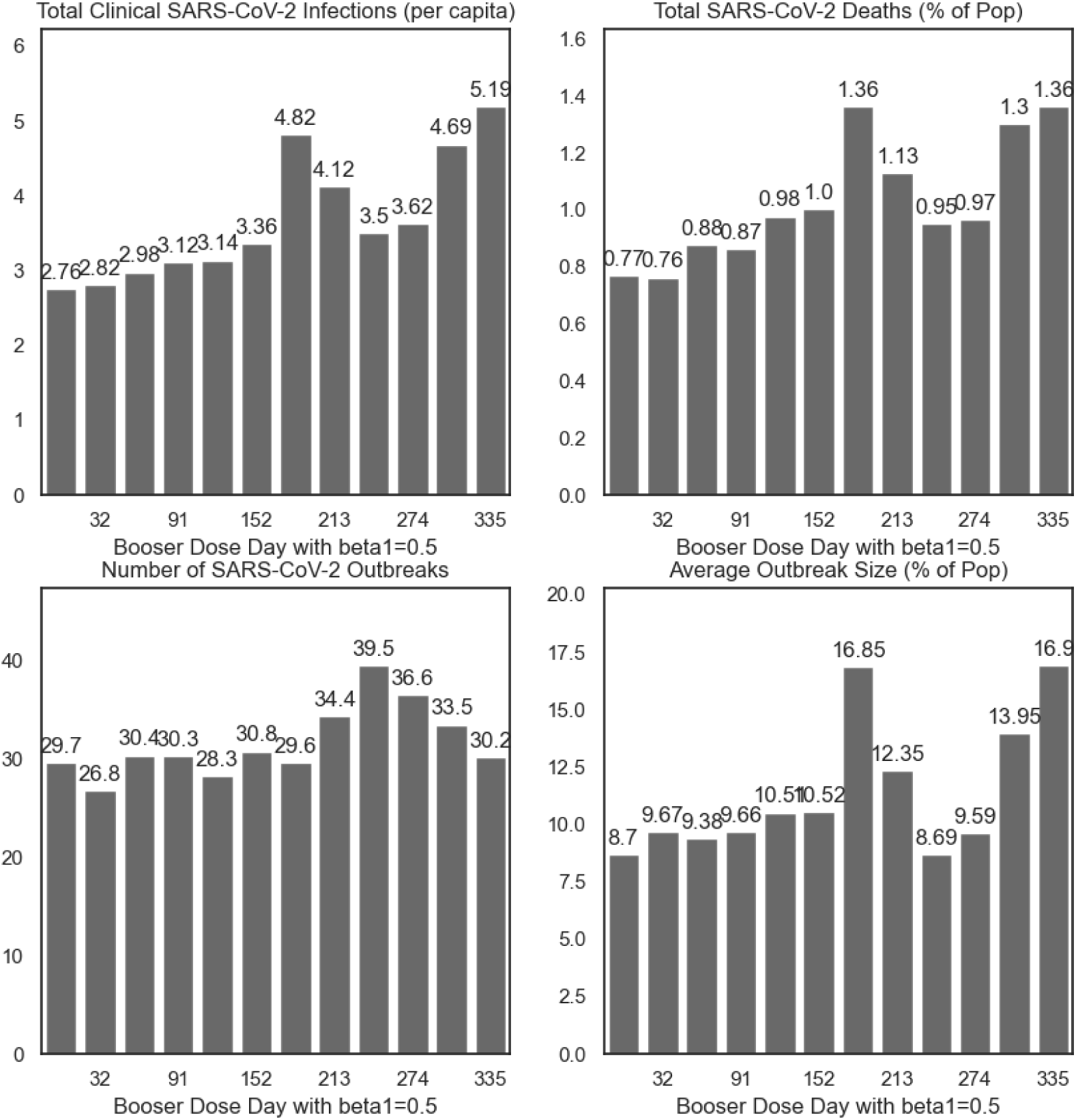
Summary of simulations by day of booster dose distribution, varied as the first of each month, in the presence of seasonal forcing of the transmission parameter *β*. Simulations are run for 10 years in replicates of 10.

**Figure 8:**
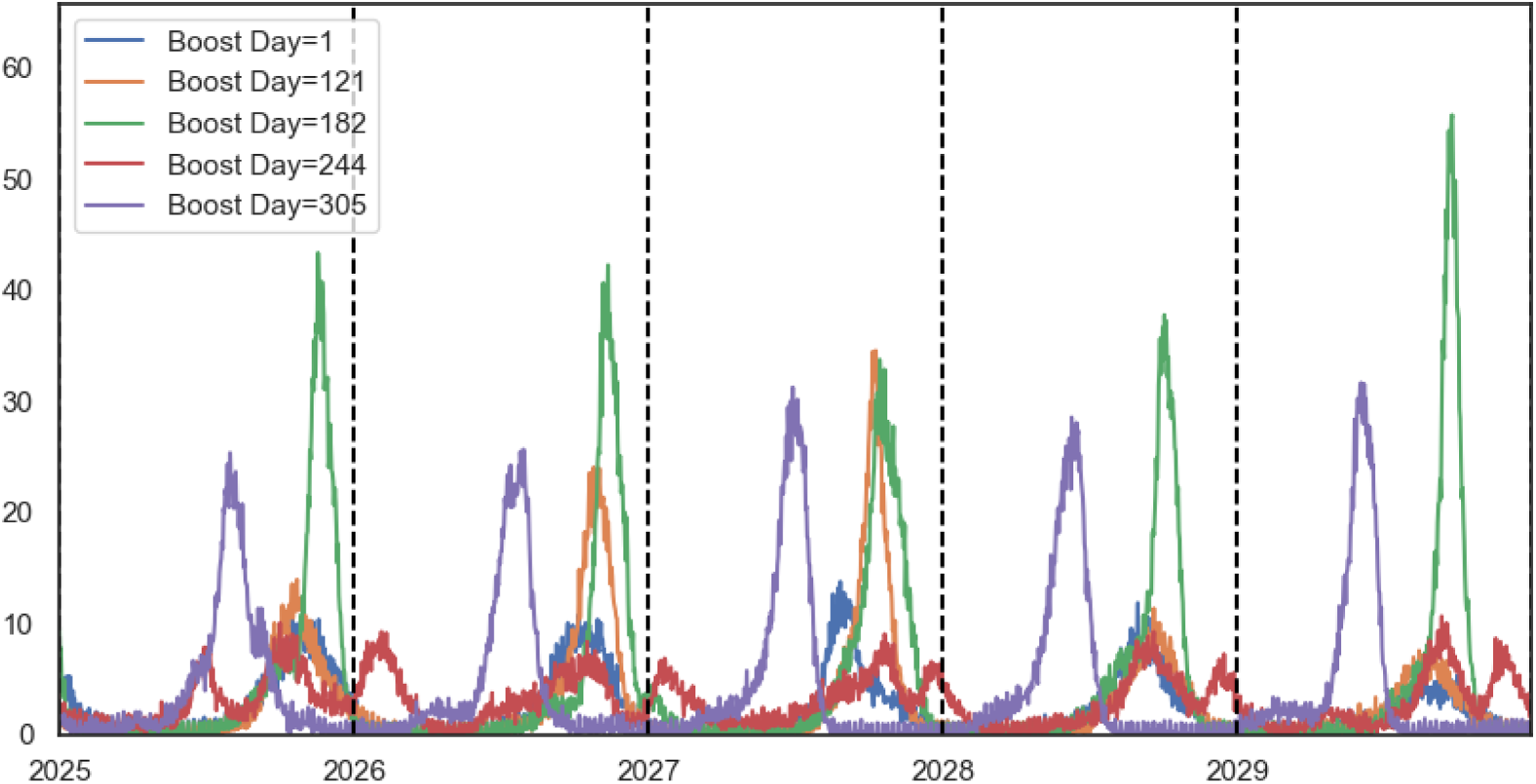
5-year trajectory of simulations by day of booster dose distribution, for selected distribution days: Jan 1st (day 1), May 1st (day 121), July 1st (day 182), Sept 1st (day 244), and Nov 1st (day 305), in the presence of seasonal forcing of the transmission parameter *β*, averaged across 10 replications.

As a sensitivity test, we varied the duration of immunity after infection and vaccination, and find that this parameter has a substantial effect on the timing and size of outbreaks. As this parameter governs the rate and which susceptibles are effectively re-introduced into the population, our results are onsistent with M. G. Baker, Peckham, and Seixas 2020. Recurrent outbreaks of SARS-CoV-2 are driven by the seasonality included in the model but also by the effect of waning natural and vaccine-derived immunity, such that even models without seasonal forcing and booster dose distribution may exhibit predictable outbreaks (figure 9). Rather than these outbreaks occurring at specific times throughout the year, they occur a certain amount of time after the previous outbreak—generally equal to the duration of complete immunity. At 6 month immunity, generally two outbreaks occur per year, spaced slightly more than 6 months apart, with periodic secondary outbreaks between; at one year of full immunity, large outbreaks occur slightly more than one year apart, without secondary outbreaks. These simulations indicate that preparations for outbreaks should include evaluation of the previous major outbreak.

**Figure 9:**
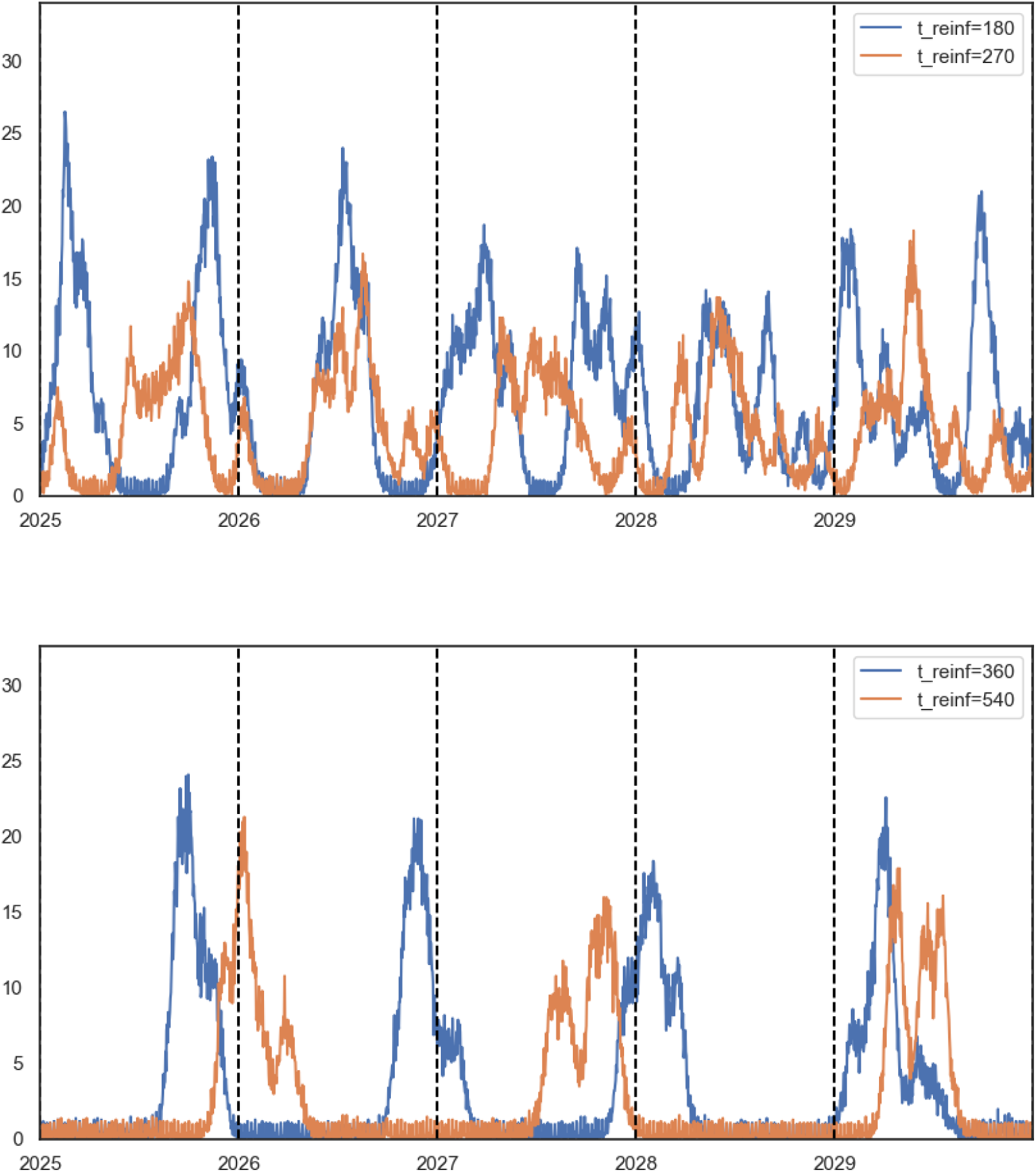
Trajectory of SARS-CoV-2 clinical infections with varied duration of complete immunity after infection and vaccination, illustrating how outbreak timing can be affected by immunity. Top: full immunity lasting for 180 days (6 months) and 270 days (9 months); bottom: full immunity lasting for 360 days (12 months) and 540 days (18 months). Averaged across 10 replications.

We also explored the possibility of isolation as a means to control SARS-CoV-2, despite it being unlikely to be used as a general control strategy in the future. Isolation remains an effective way to limit the spread of SARS-CoV-2 in the steady state, however only the higher degrees of isolation— reducing non-household contacts among clinically infectious individuals to less than 50% of their normal amounts as compared to a ‘business as usual’ scenario— has a substantial effect on transmission dynamics (figure 10). At 50% isolation, an average of 2.72 clinical infections occur per capita, down from 4.84 when clinically infectious nodes have 75% of their normal random contacts and 6.22 in the complete absence of isolation. With 50% isolation, the outbreak size drops dramatically to 6.05% of the population infected during an average of 40.8 outbreaks, down from from 18.7% of population infected in an average of 32.2 outbreaks in the absence of isolation. More extreme isolation reduces the severity of outbreaks even further: at 25% of normal contacts, clinical infections average one per capita; with perfect isolation, clinical infections are less than 0.5 per capita.

**Figure 10:**
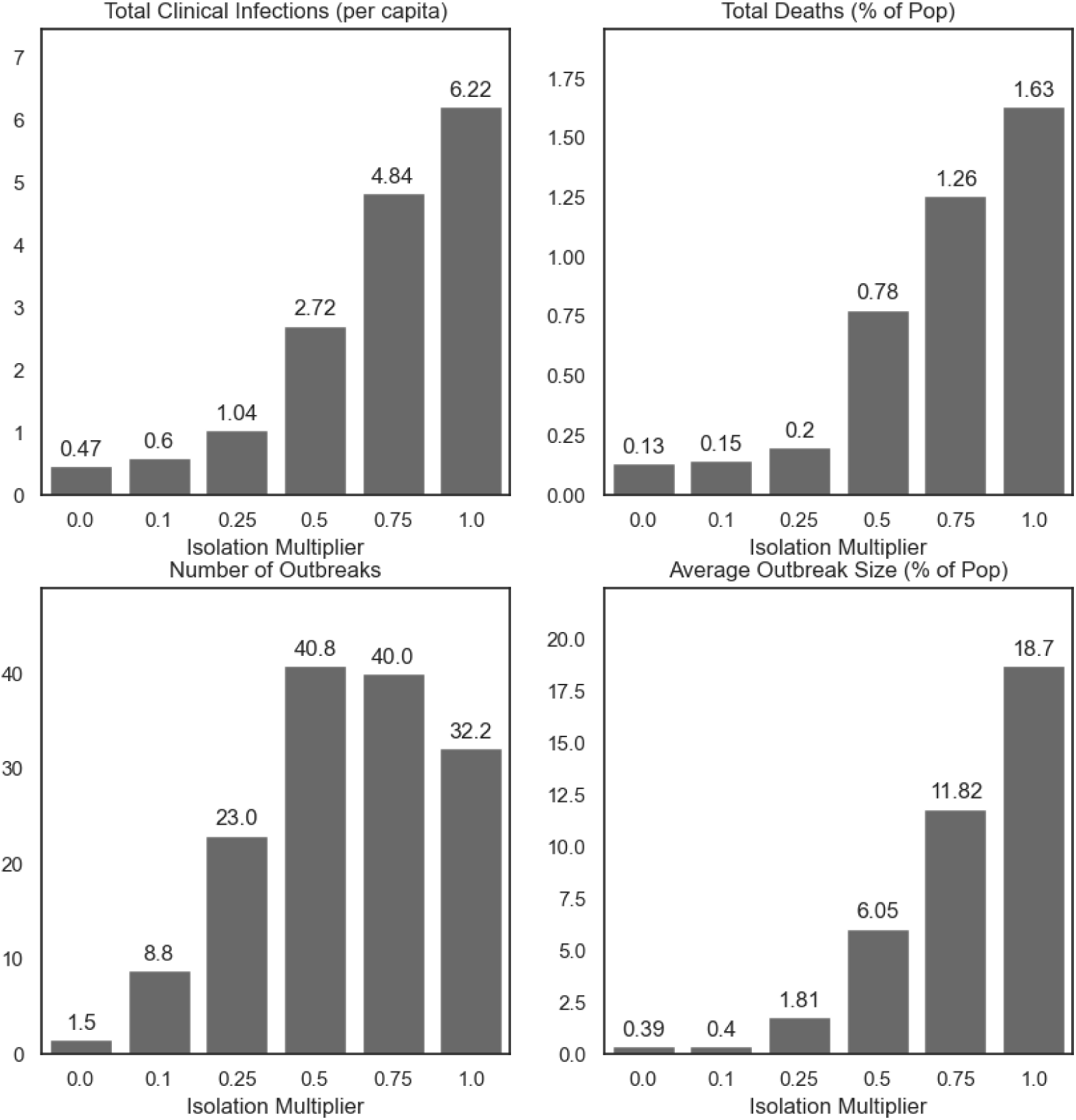
Summary of SARS-CoV-2 clinical infections, deaths, number of outbreaks, and average outbreak size for different levels of isolation, in the absence of seasonal forcing or vaccine distribution. Isolation multiplier is a factor used to scale a clinically infectious node’s random, non-household contacts; 1.0 indicates business as usual and 0 indicates perfect isolation.

## 4 Discussion

Across all simulations, we observed frequent and predictable SARS-CoV-2 outbreaks over a 10-year period, even with the annual distribution of booster doses as the primary disease-averting intervention. Depending on the epidemiology of the pathogen—namely, should SARS-CoV-2 exhibit seasonality in transmission probability and contact—outbreaks may occur at predictable times of the year, and distribution of booster doses may be able to mitigate the worst of seasonal outbreaks. Our results are consistent with R. E. Baker et al. 2020, who find that outbreak cycles are primarily determined by the levels of susceptibility in the population, although seasonality is an important moderator in outbreak dynamics. Different vaccination campaigns may be needed in areas that exhibit stronger transmission seasonality. We find that distributing booster doses in the first half of the year—January through May—may be an effective strategy at limiting recurrent outbreaks depending on the seasonality exhibited by the pathogen. We find that in simulations without seasonal forcing, distributing booster doses in the first half of the year is most effective at limiting outbreaks; however, with the inclusion of seasonal forcing of transmission and contact, distributing booster doses in September or October may limit the average outbreak size the most. In these simulations, distributing booster doses in the late fall ‘shifts’ the outbreak to the summer, when transmissibility is lower. Since influenza vaccines are distributed in the fall, including booster doses for SARS-CoV-2 at the same time may be easiest in implementation, but less successful than Springtime distribution in limiting outbreaks should

### SARS-CoV-2 fail to exhibit transmission seasonality

In addition to illustrating how vaccination interventions can avert the burden of illness and death due to SARS-CoV-2, our simulations further understanding of how to prepare for future SARS-CoV-2 outbreaks. Although distributing booster doses in early fall—like annual vaccinations for influenza—avoids a large wintertime outbreak in a seasonally forced environment, multiple smaller outbreaks may occur throughout the year. This may be preferable to avoid exceeding the treatment capacity of the health system. However, this strategy may not be effective in a less seasonally-variable climate, where booster dose timing merely shifts the main outbreak to later in the year after immunity wanes. Overall, we generally observe variance in mortality proportional to the number of clinical infections that occur in the simulations, indicating both that the most effective strategies for limiting clinical infections also limit deaths. Although we focus on the number of infections as the primary outcome, reducing the number of deaths due to SARS-CoV-2 may be possible with the interventional strategies outlined here.

While agent-based models have been used in a variety of applications for COVID-19, ours represents one of the first to examine long-term dynamics using real population contact data. Our results hint at a rather bleak outlook for the future of SARS-CoV-2: that outbreaks are extremely likely to continue given that natural and vaccine-derived immunity wanes over time. However, annual booster doses—especially when timed properly—and isolation of infectious cases may be effective control strategies. The investigation above tells us that outbreaks can be expected to occur more frequently than annually, and the epidemiology of the pathogen—namely, the base transmission probability and the duration of immunity after infection and vaccination— determines the frequency and severity of outbreaks more than vaccination timing and within a reasonable range of effectiveness. As our approach to SARS-CoV-2 shifts from eradication to management, the strategies presented here show how we can time the distribution of doses to minimize strain on the healthcare system and limit chronic complications from SARS-CoV-2 infections.

Our model has a number of shortcomings that limit its generalizability. First, we are limited by computing resources to a population of 1000 households (approximately 3, 200 individuals). During development, we found that using too few households resulted in simulations that were highly unstable; with a larger population, simulation results were more consistent between runs but took much longer to complete. As a result, we chose the number of households to balance numerical stability while maintaining a reasonable run time of approximately 4 minutes per simulation. Larger populations may exhibit different dynamics as the spread of infection may take longer; as such, it is not known presently if this chosen population size is representative of a larger population or is limited in generalizability to smaller communities. As well, like Roubenoff, Feehan, and Mahmud (2023), we borrow contact patterns for the youngest age group from the POLYMOD UK survey (Mossong et al. 2008), which may not be representative of that age group in the US during the COVID-19 pandemic. However, since our application here is in consideration of the long-term patterns of SARS-CoV-2,’ we believe that these contacts represent a return to ‘business as usual’ for school children.

A key feature of our model is in the random network generation, that produces daily draws of random contacts according to the network configuration model parameterized with degree and duration from the BICS survey. Like any random network generation, the network structure of contacts produced by this model may affect transmission dynamics. Future work should draw inspiration from the COVASIM agent based model (Kerr et al. 2021), which allows for comparison of outbreak dynamics between simulations with Poisson random networks, Hybrid networks, or SynthPops networks depending on the needs of the simulation and data available. As well, we do not include any assortative mixing as emphasized by the model used by Roubenoff, Feehan, and Mahmud 2023 or serial (repeat) contacts, instead choosing a network generation function that maintains the degree of each node. Employment contacts, like school contacts for children under the age of 18, provide a consistent set of individuals having contact most days; inclusion of these such contacts may affect outbreak in unpredictable ways. Future development of the model should include associativity by age and employment status; however, doing so was outside of the scope of the present analysis.

## Implementation Overview

The BICS ABM model is implemented in the `C++` language and with use of the `iGraph`-C library (G. Cśardi and Nepusz, T. 2006; Gábor Cśardi et al. 2023), and a Python 3.8 API. The core C++ implementation was chosen over other languages, like `Python` or `R`, to maximize speed. Full implementation details are given in the supplementary material. Each simulation of 1000 households for 10 years completes in approximately 4 minutes on an Apple MacBook Air M1, and the entire suite of simulations completes in approximately 8 hours. Replication code is publicly available at https://github.com/eroubenoff/BICS_ABM.

## Data Availability

All data produced in the present study are available upon reasonable request to the authors

## 5 Model Supplement

The model described here is a stochastic Agent-Based Network Simulation for COVID-19 transmission that utilizes data collected as part of the Berkeley Interpersonal Contact Survey to simulation population structure and contact networks. The core algorithm is written in `C++`, compiled using Apple Clang++17, and utilizes CMake 3.16 and utilizes the `igraph 10.4` library (G. Cśardi and Nepusz, T. 2006; Gábor Cśardi et al. 2023), and includes a Python 3.8 user API. The BICS ABM C++ library is compiled to a dynamic library for linkage to the Python API and can be used with an external C++ program.

The BICS ABM C++ library hinges on two input data structures and their Python equivalents: `Params`, a C-struct that contains the input parameters, documented in table 1; and an array of the simulation population. The input population requires a strict format: each row is an individual and each column represents individual-level data passed to the simulation, flattened to one dimensional array of floats in column-major (Fortran-style) order. Currently, 8 fields are required for each simulated node: the node’s household id; age, represented as a categorical index 0-8; gender, where 0 corresponds to male and 1 corresponds to female; number of non-household contacts; number of school contacts, which is taken to be zero for adults; number of times left home (unused presently, but maintained for legacy purposes); vaccine priority (see 5.1.5); and a boolean indicating if the node uses NPIs or not. Parameters must be supplied indicating the dimensionality of the dataset. The simulation returns a `trajectory`, a C-struct containing an hourly time-series of all disease states.

The Python API contains a class `BICS ABM`, which is a wrapper around all of the above utilizing the `ctypes` library for cross-language functionality. Parameters can be passed to the model through the Python API identically as to the C++ library and we recommend interacting with the simulation through the Python API. The class constructor for `BICS ABM` takes any of the arguments to `Params`, runs the simulation, and saves as fields the components of the resultant trajectory as a `numpy.ndarray`; as such, the trajectory of clinical cases can be accessed as `BICS ABM.Cc`. The population can be accessed through `BICS ABM. pop`.

**Table 1:**
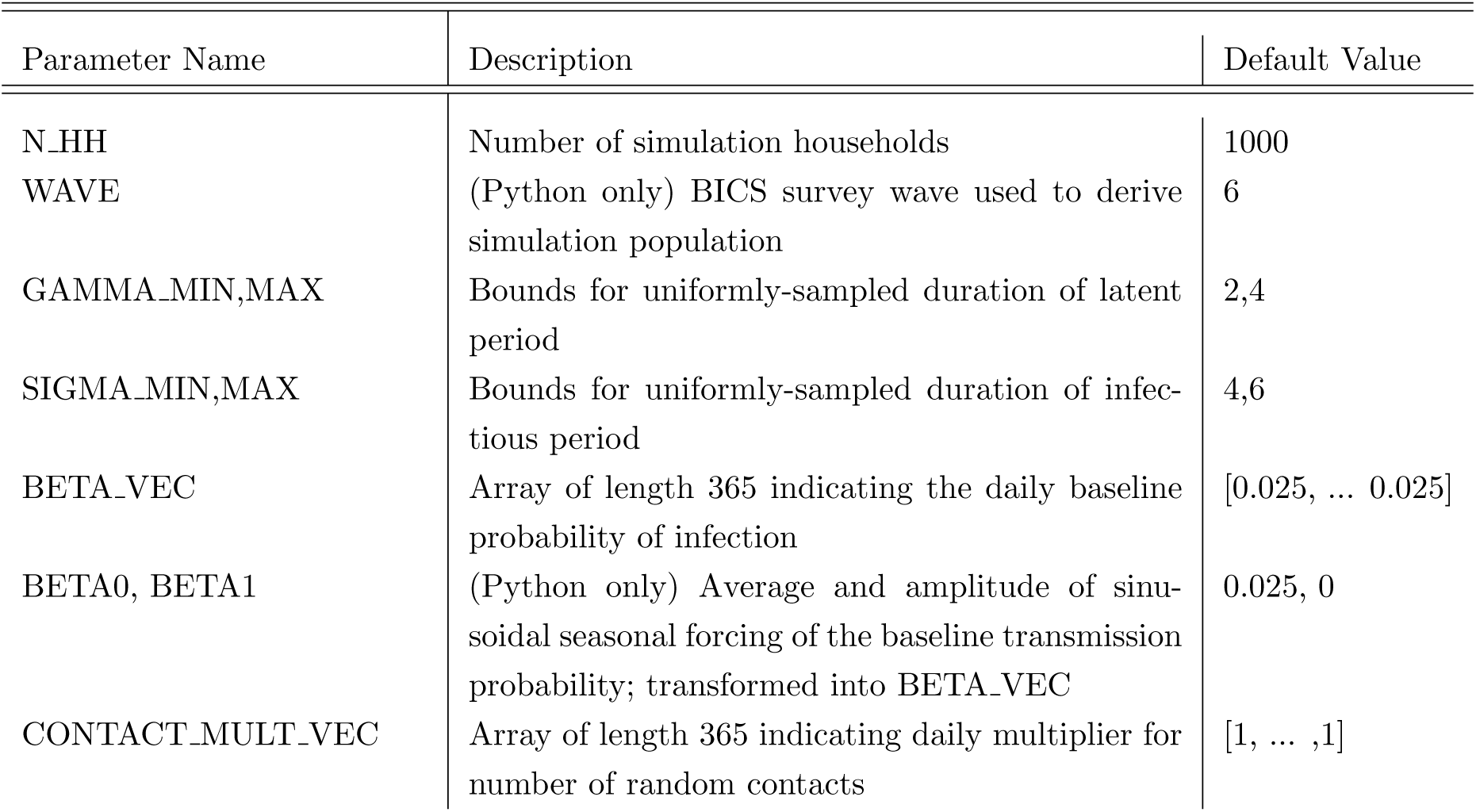
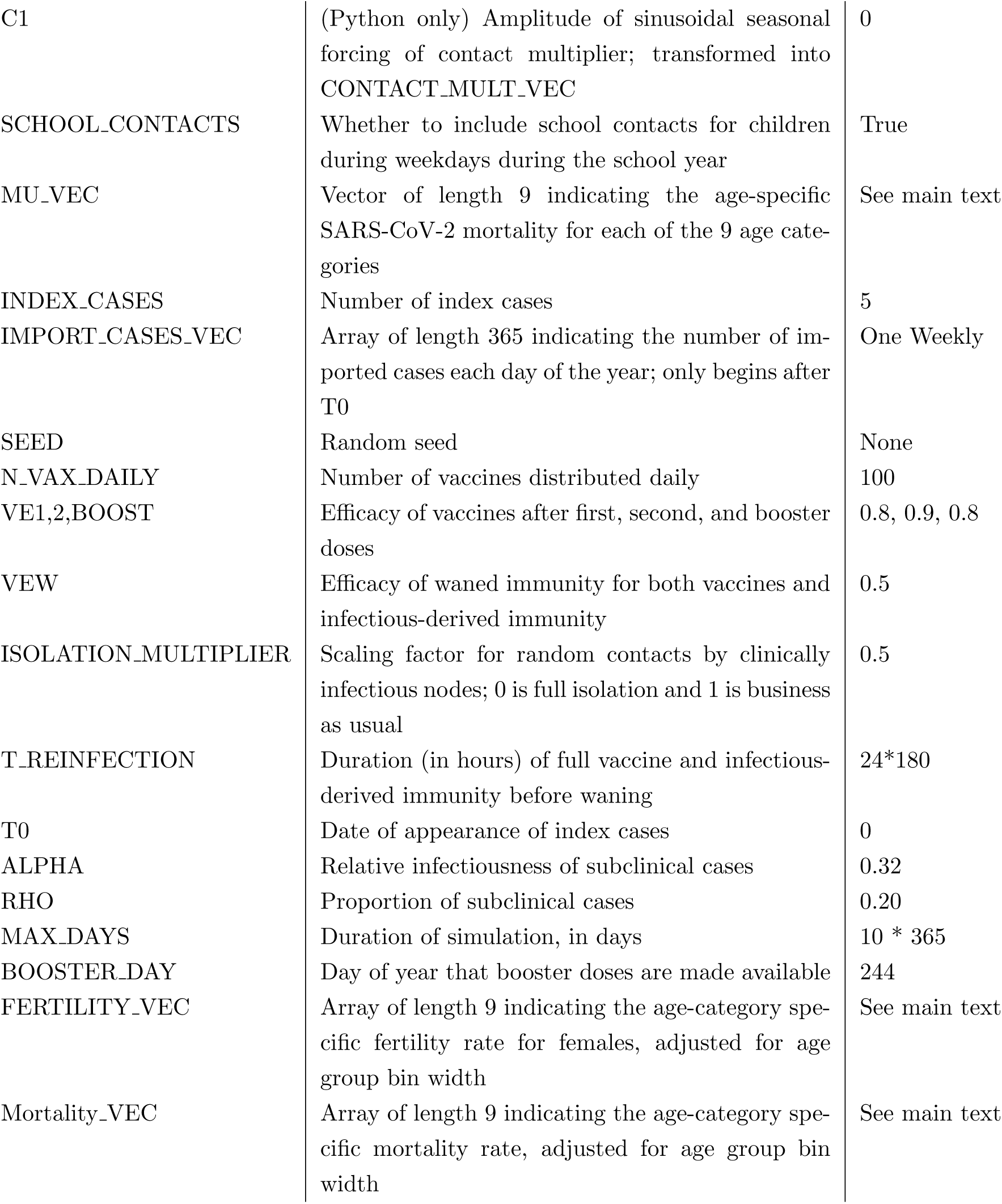
Parameters used in the ABM model.

| Parameter Name | Description | Default Value |
| --- | --- | --- |
| N_HH | Number of simulation households | 1000 |
| WAVE | (Python only) BICS survey wave used to derive simulation population | 6 |
| GAMMA_MIN,MAX | Bounds for uniformly-sampled duration of latent period | 2,4 |
| SIGMA_MIN,MAX | Bounds for uniformly-sampled duration of infectious period | 4,6 |
| BETA_VEC | Array of length 365 indicating the daily baseline probability of infection | [0.025, ... 0.025] |
| BETA0, BETA1 | (Python only) Average and amplitude of sinusoidal seasonal forcing of the baseline transmission probability; transformed into BETA_VEC | 0.025, 0 |
| CONTACT_MULT_VEC | Array of length 365 indicating daily multiplier for number of random contacts | [1, ... ,1] |
| C1 | (Python only) Amplitude of sinusoidal seasonal forcing of contact multiplier; transformed into CONTACT_MULT_VEC | 0 |
| SCHOOL_CONTACTS | Whether to include school contacts for children during weekdays during the school year | True |
| MU_VEC | Vector of length 9 indicating the age-specific SARS-CoV-2 mortality for each of the 9 age categories | See main text |
| INDEX_CASES | Number of index cases | 5 |
| IMPORT_CASES_VEC | Array of length 365 indicating the number of imported cases each day of the year; only begins after T0 | One Weekly |
| SEED | Random seed | None |
| N_VAX_DAILY | Number of vaccines distributed daily | 100 |
| VE1,2,BOOST | Efficacy of vaccines after first, second, and booster doses | 0.8, 0.9, 0.8 |
| VEW | Efficacy of waned immunity for both vaccines and infectious-derived immunity | 0.5 |
| ISOLATION_MULTIPLIER | Scaling factor for random contacts by clinically infectious nodes; 0 is full isolation and 1 is business as usual | 0.5 |
| T_REINFECTION | Duration (in hours) of full vaccine and infectious-derived immunity before waning | 24*180 |
| T0 | Date of appearance of index cases | 0 |
| ALPHA | Relative infectiousness of subclinical cases | 0.32 |
| RHO | Proportion of subclinical cases | 0.20 |
| MAX_DAYS | Duration of simulation, in days | 10 * 365 |
| BOOSTER_DAY | Day of year that booster doses are made available | 244 |
| FERTILITY_VEC | Array of length 9 indicating the age-category specific fertility rate for females, adjusted for age group bin width | See main text |
| Mortality_VEC | Array of length 9 indicating the age-category specific mortality rate, adjusted for age group bin width | See main text |

### 5.1 Model Pseudocode

Psuedocode for the model is given below:

1. In Python API: Establish the following parameters governing the population structure: number of households *n_hh_*, survey wave, and vaccine priority. Establish all other transmission parameters and store them in an object `params`.
2. Generate *n_hh_*households from the corresponding survey wave using the procedure described below. Assign each household a unique household identifier.

(a) First, the distribution of adult ages and sex is derived from ACS data, and a set of *N HH* households are sampled from this distribution.
(b) A ‘head’ of household is randomly chosen from the BICS survey data. A respondent is eligible to be head of household *h_i_* if they are the corresponding age and sex for the sampled household head. Eligible household heads are chosen with replacement and with probability adjusted for survey weights.
(c) Finally, households are filled by sampling (with replacement and adjustment for survey weights) from the set of BICS respondents who mach each of *h_i_*’s reported household members’ age, gender, and household size. Children under 18 are not ascertained in the survey; children are instead sampled from the POLYMOD survey. The max size of a household is 6 as as respondents were only asked to report 6 of their household members.
3. Assign vaccine priority to all nodes in the network based off of the rules provided as input, as elaborated in section 5.1.5.
4. Create and pass the `params` and population object to the C++ core algorithm.
5. In C++ core: Determine the household contact network assuming that all nodes have contact with all members of their household. Randomly draw a school contact network for children under the age of 18.
6. Repeat the following procedure representing one ‘day’ of simulation time, where each day contains 24 ‘hours’ of simulation time, until either no nodes are exposed or infectious OR the simulation has occurred for a supplied maximum number of days.

(a) If hour == 0 (midnight) and an index case is supplied to appear on the current simulation day, transition one node at random into ‘Exposed’ status.
(b) Each hour between midnight and 8am: assume that all nodes have contact with all members of their household. Execute `transmission` and `decrement` procedures for all nodes.
(c) 8am: Distribute vaccine doses to `n vax` nodes awaiting any dose of the vaccine.
(d) 8am: generate a random graph of daily contacts using the procedure outlined in section 5.1.3; assign a random duration and start time for all contacts.
(e) Each hour between 8am and 6pm: connect all random contacts; disconnect each node having a random contact from their household nodes; `transmit`; and `decrement`. Reconnect nodes after termination of random contact.
(f) Each hour between 6pm and midnight: transmit within households assuming that all nodes have contact with all members of their household. Execute `transmit` and `decrement` procedures.
7. At the conclusion of the simulation, return trajectories of each disease and vaccination status.

#### 5.1.1 Update Handlers

The C++ program contains a centralized method for handling and dispatching changes to nodes, edges, and the graph itself. Classes exist for five types of changes can be executed: `UpdateGraphAttribute`, `CreateEdge`, `DeleteEdge`, `UpdateEdgeAttribute`, and `UpdateVertexAttribute`. Vertices cannot be created or destroyed using the update handler. All updates are stored in wrapper class `UpdateList`, which contains an overloaded method `UpdateList.add update()` to add an update of each type. Updates are then dispatched with the method `UpdateList.add updates to graph(igraph t*)`, which takes a reference to the graph object as an argument to perform the updates.

The centralized update handlers were developed to streamline the attribute interface. Although the `igraph` API contains methods for updating individual node or edge attributes, it is more computationally efficient to pull all of the attributes, make all changes, then push them back to the graph. Since the `igraph` API involves many dynamically-allocated objects, this meant keeping track of many pointers, being sure to free all used vectors of attributes. With a project of this size, adding a centralized way of dispatching updates helped with debugging many issues with memory management, at the cost of a slight function overhead. As well, the object-oriented interface allows for saving of all, say, household edges in a single object to be connected and disconnected as needed.

#### 5.1.2 Decrement procedure

The `decrement` procedure is among the most important function in the C++ core for tracking the progression of nodes through simulation time. Each ‘event’ that can occur to a node (infection, development of clinical or subclinical infectiousness, recovery, waning immunity, becoming vaccinated, etc) is accompanied by a duration of the event. The `decrement` procedure decrements the time remaining at each status, and for some events (for example, recovery from infectiousness) will automatically trigger a status change; for other events (like eligibility for vaccination), eligible nodes are placed in a queue for the next event to occur.

#### 5.1.3 Random contacts

Random contacts are drawn for the daytime hours, 8am-6pm, using the network configuration model (as described in the main text; Albert-ászó Barabási 2021). First, the number of daily-nonhousehold contacts is supplied for each simulation node; this number is first multiplied by the isolation multiplier if the node is clinically infectious and contact multiplier if included in the model, then taken to be a random draw from a Poisson distribution with rate parameter equal to the product of all three terms. This is done to allow for minor stochasticity in the model. Each node is assigned a number of stubs equal to this Poisson random draw; if the total number of stubs subs to an odd number, one stub is randomly deleted until the sum is an even number. Should this sum be zero stubs then the procedure aborts. The configuration model is drawn using the `igraph degree sequence game` from the `igraph` library; the graph is then simplified to remove self-edges but not multi-edges.

#### 5.1.4 Non-Pharmaceutical Interventions

We include a single generic Non-Pharmaceutical Intervention (NPI) intended to capture the combined transmission-preventing power of mask usage, gloves, and physical distancing. In wave 6, about 60% BICS respondents reported the usage of any of these possible NPIs in any of their reported contacts; any simulation nodes representing these respondents are given an NPI status of `True`. During the daytime simulation

**Figure 11:**
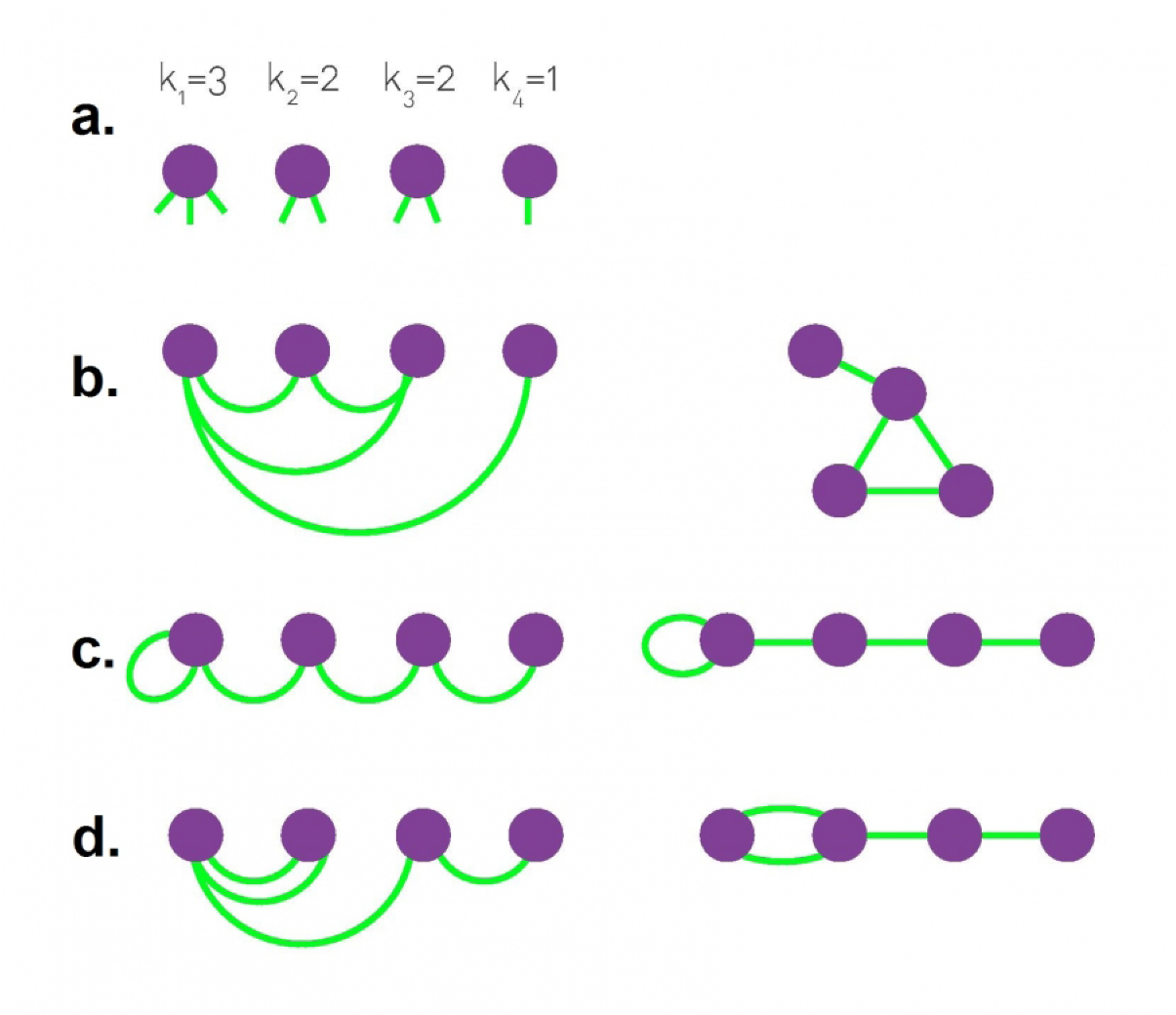
Albert-ászó Barabási 2021’s diagram representing realizations of the Network Configuration Model showing multiple ways of connecting the four nodes in panel (a) with corresponding degree *k*. (b): no self- or multi-edges; (c): allowing self-edges but not multi-edges; (d): allowing multi-edges, but not self-edges. Our application would allow for configuration (d) but not (c). procedure, if two non-household nodes are connected who both have an NPI status of `True`, then their transmission probability is lowered by a supplied parameter representing the strength of these NPIs (see below).

#### 5.1.5 Vaccine Distribution

Nodes in the population are assigned a discrete priority level for vaccination before the simulation begins. A set number of vaccines are distributed daily among nodes with the highest priority level until no nodes in that priority level remain; remaining doses are distributed among nodes with the next highest priority level, repeating until the day’s number of vaccines are exhausted. A priority level of *−*1 indicates that the node declines or is ineligible for vaccination. Nodes are eligible to receive the second dose of the vaccine 25 days after they received first dose. After a period of time, vaccine efficacy is assumed to wane; at this point, nodes are eligible to receive an additional booster dose.

#### 5.1.6 Demographic Rates

Fertility and all-cause mortality are incorporated in the simulation according to published age-specific rates, aggregated for each age group in the simulation:

**Figure 12:**
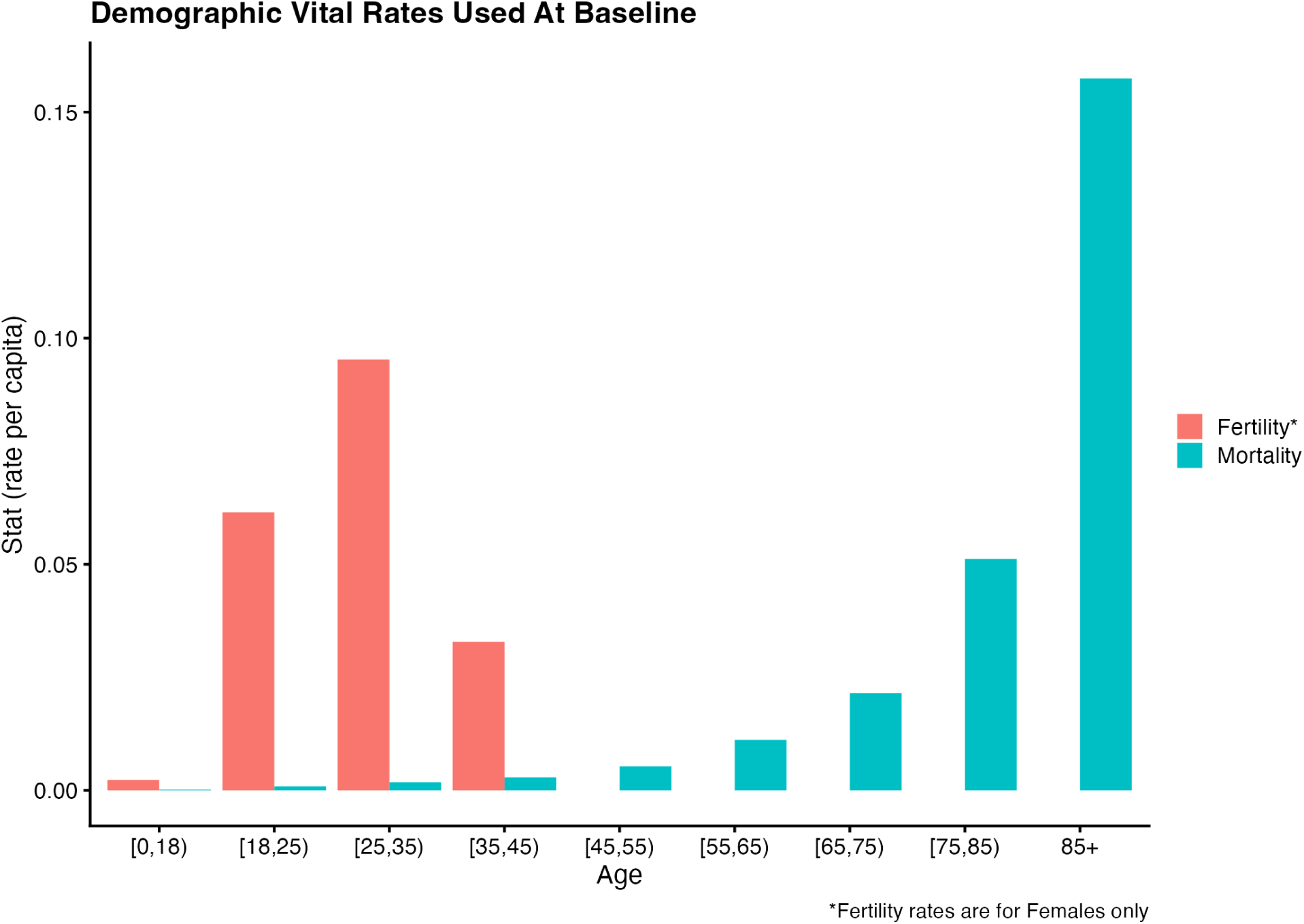
Baseline demographic vital rates used in the simulation.

### 5.2 Supplementary Figures

#### 5.2.1 Booster Dose Distribution Day, High Uptake

**Figure 13:**
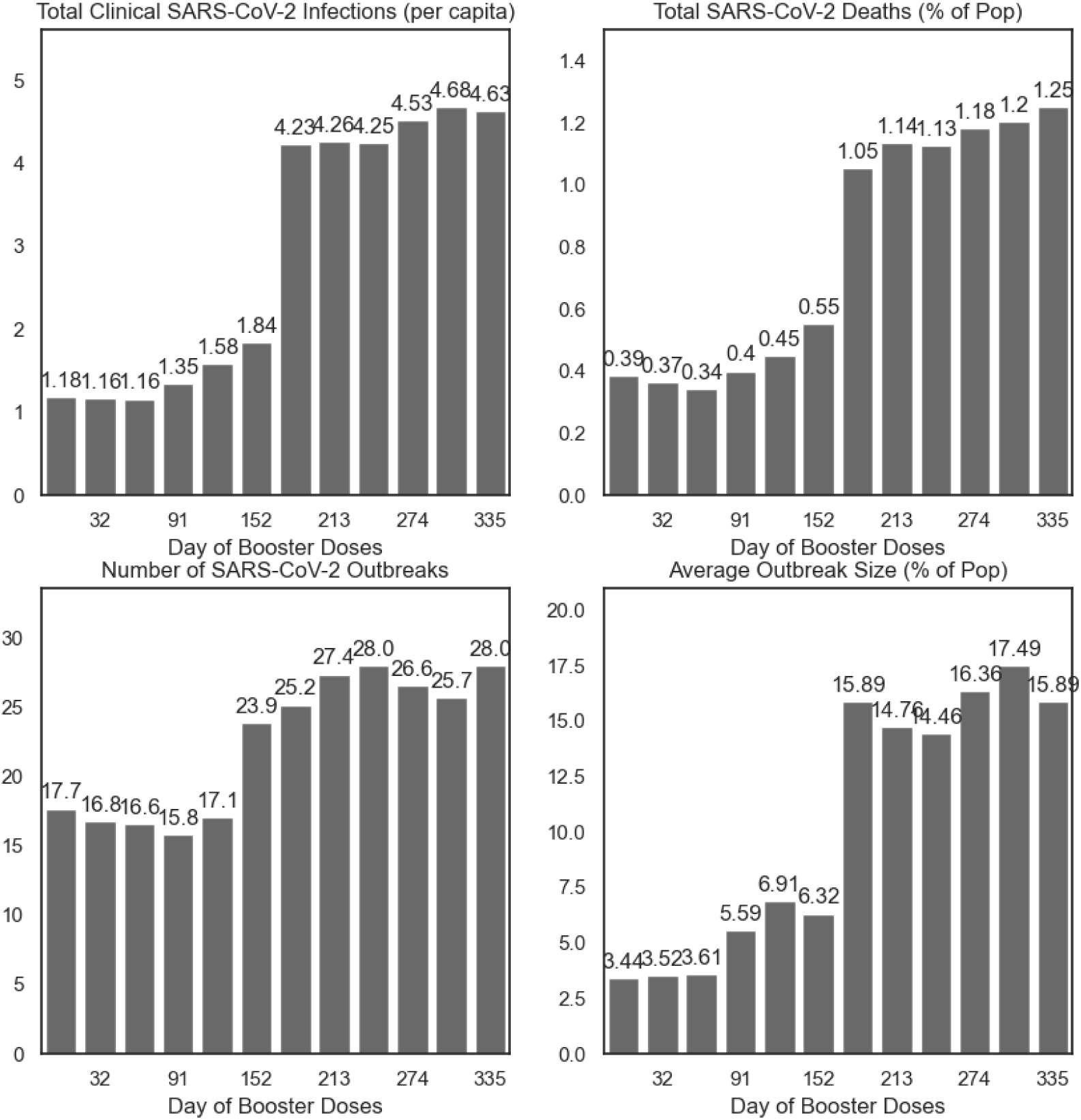
Summary of simulations by day of booster dose distribution, varied as the first of each month, in the absence of seasonal forcing of the transmission parameter *β*, with 90% vaccine uptake.

**Figure 14:**
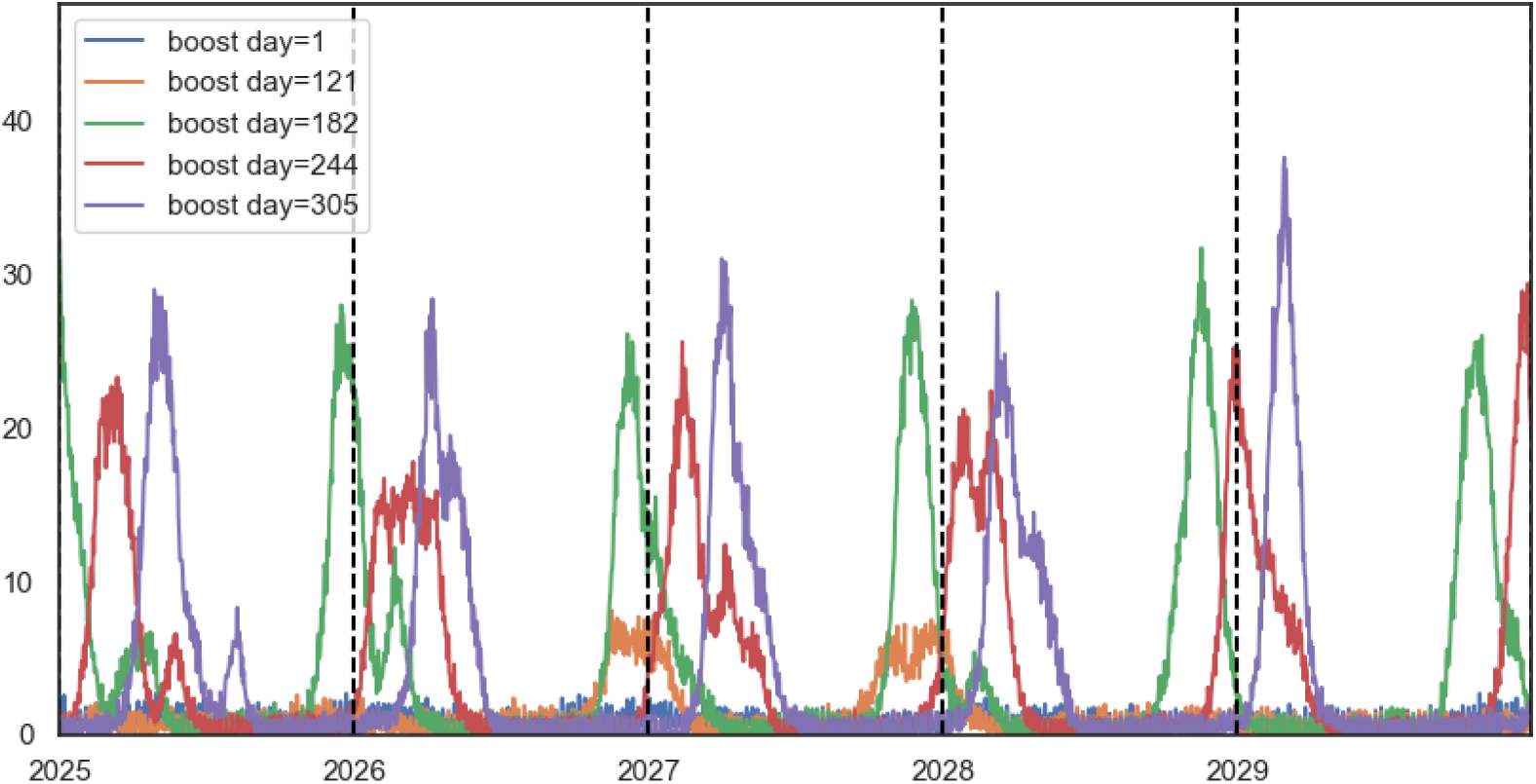
Trajectory of simulations by day of booster dose distribution, without seasonal forcing, for selected distribution days: Jan 1st (day 1), May 1st (day 121), July 1st (day 182), Sept 1st (day 244), and Nov 1st (day 305), with 90% uptake.

#### 5.2.2 Seasonality in contact rates

**Figure 15:**
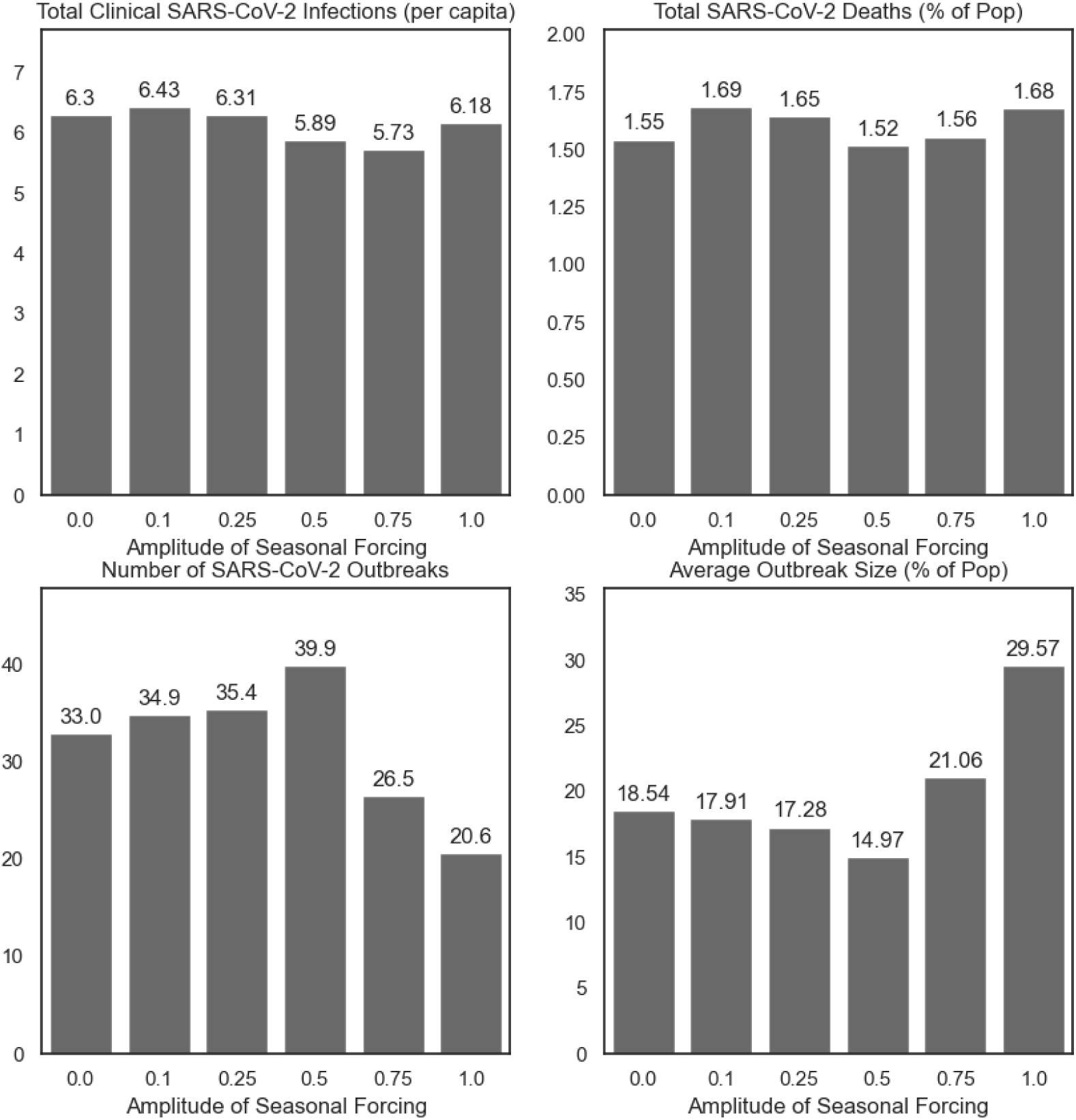
Summary of simulations at selected levels of *c*_1_, the amplitude of seasonal forcing of the contact parameter.

**Figure 16:**
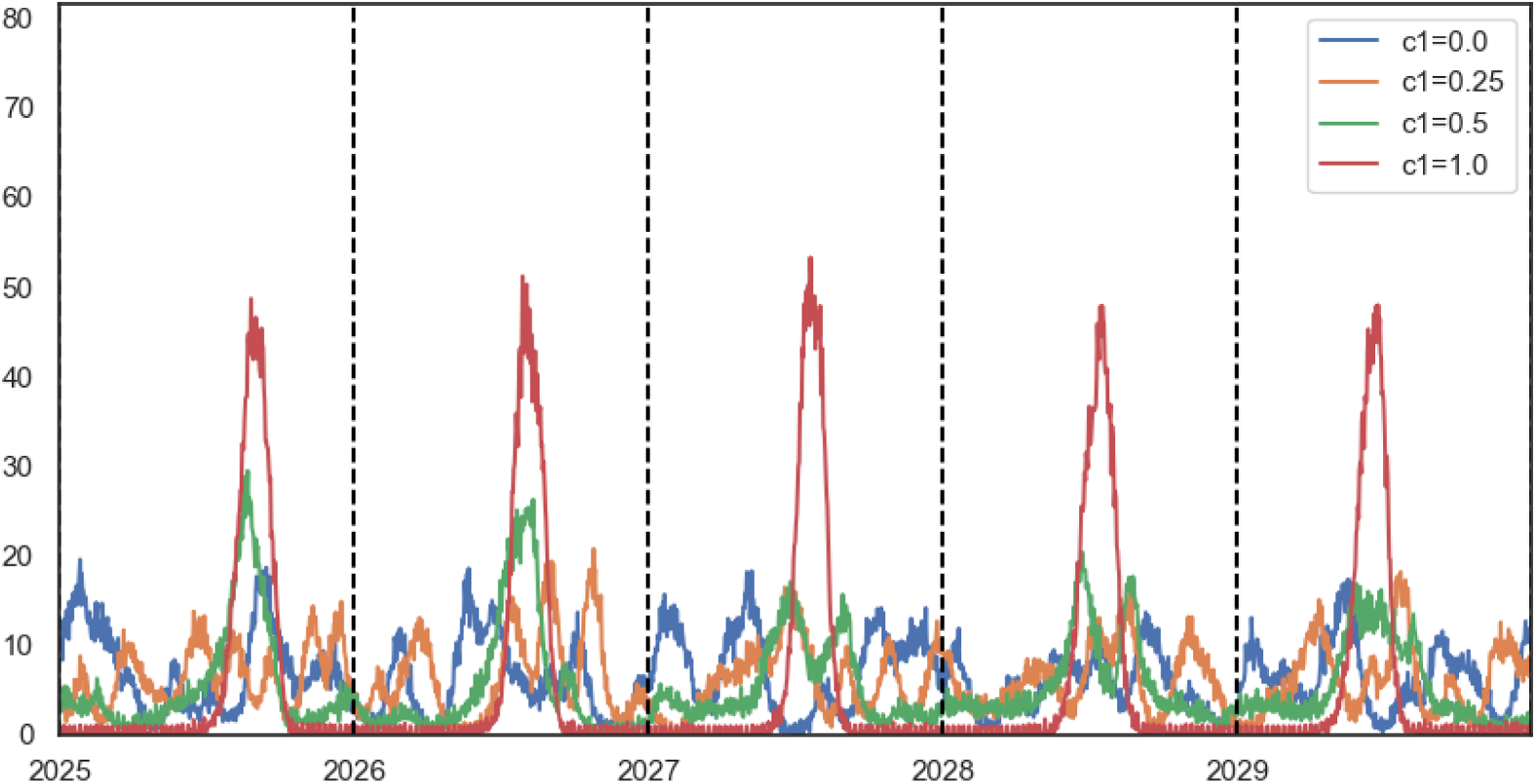
Trajectories at selected levels of *c*_1_, the amplitude of seasonal forcing of the contact parameter.

#### 5.2.3 Isolation of infectious cases

**Figure 17:**
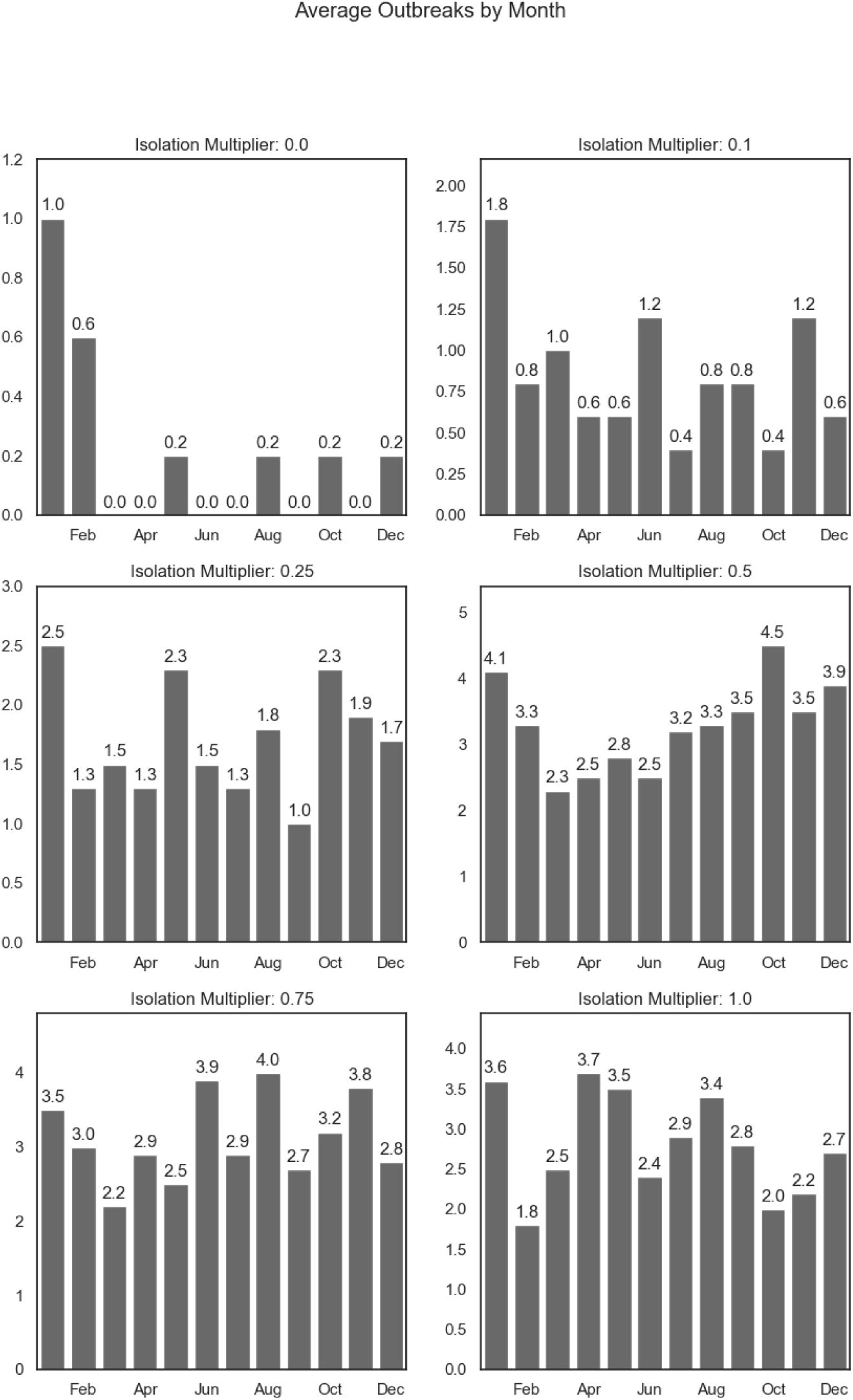
Outbreak seasonality at selected levels of isolation

**Figure 18:**
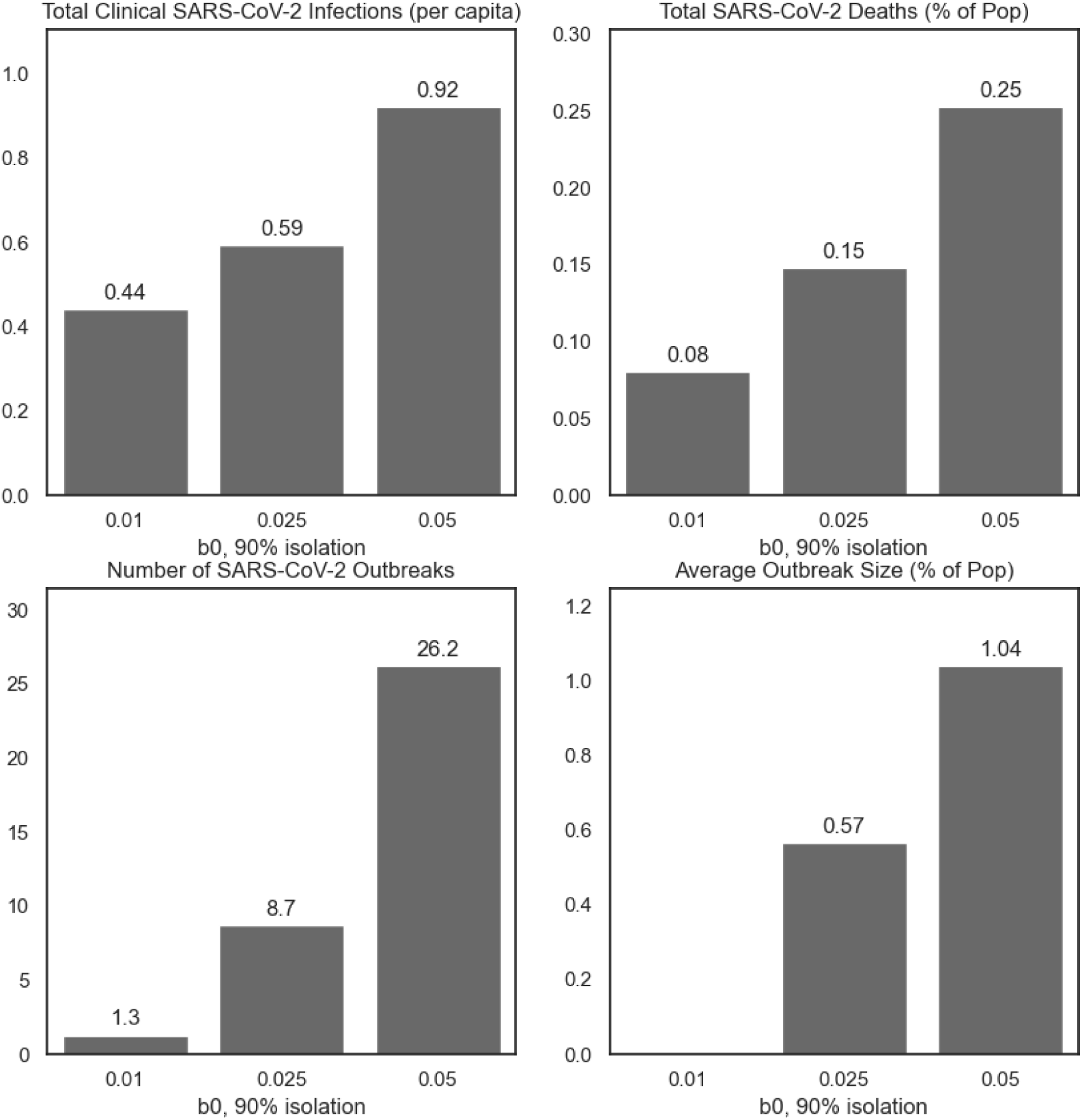
Summary of simulations at selected levels of *β*_0_ with 90% isolation of clinically infectious cases (corresponding to an isolation multiplier of 10%).

## Footnotes

1 Some fields collected as part of the BICS survey are not available in the POLYMOD data and are imputed. For the present application, only mask/NPI usage is unavailable, and is taken to be the False.

2 See Weaver et al’s review of the following studies, among others: Chin et al. 2020; J. Liu et al. 2020; Ma et al. 2021; Matson et al. 2020; Morris et al. 2021; Nottmeyer and Sera 2021; Raiteux et al. 2021; Riddell et al. 2020; Sera et al. 2021; Smith et al. 2021; Wu et al. 2020

## References

Ajelli, Marco et al. (June 2010). “Comparing large-scale computational approaches to epidemic modeling: Agent-based versus structured metapopulation models”. In: BMC Infectious Diseases 10.1, p. 190. issn: 1471-2334. doi: 10.1186/1471-2334-10-190. url: 10.1186/1471-2334-10-190 (visited on 09/01/2021).

Albert-László Barabási (2021). Network Science by Albert-László Barabási. url: http://networksciencebook.com/ (visited on 10/06/2021).

Altizer, Sonia, et al. (Apr. 2006). “Seasonality and the dynamics of infectious diseases”. eng. In: Ecology Letters 9.4, pp. 467–484. issn: 1461-0248. doi: 10.1111/j.1461-0248.2005.00879.x.

Arduin, Héléne, et al. (June 2017). “An agent-based model simulation of influenza interactions at the host level: insight into the influenza-related burden of pneumococcal infections”. en. In: BMC Infectious Diseases 17.1, p. 382. issn: 1471-2334. doi: 10.1186/s12879-017-2464-z. url: 10.1186/s12879-017-2464-z (visited on 06/05/2023).

Baker, Jack D., et al. (Dec. 2013). “An evaluation of the accuracy of small-area demographic estimates of population at risk and its effect on prevalence statistics”. In: Population Health Metrics 11.1, p. 24. issn: 1478-7954. doi: 10.1186/1478-7954-11-24. url: 10.1186/1478-7954-11-24 (visited on 04/01/2020).

Baker, Marissa G., Trevor K. Peckham, and Noah S. Seixas (2020). “Estimating the burden of United States workers exposed to infection or disease: A key factor in containing risk of COVID-19 infection”. eng. In: PloS One 15.4, e0232452. issn: 1932-6203. doi: 10.1371/journal.pone.0232452.

Baker, Rachel E., et al. (July 2020). “Susceptible supply limits the role of climate in the early SARS-CoV-2 pandemic”. In: Science 369.6501. Publisher: American Association for the Advancement of Science, pp. 315–319. doi: 10.1126/science.abc2535. url: https://www.science.org/doi/10.1126/science.abc2535 (visited on 08/01/2023).

Bansal, Shweta, Bryan T Grenfell, and Lauren Ancel Meyers (Oct. 2007). “When individual behaviour matters: homogeneous and network models in epidemiology”. In: Journal of the Royal Society Interface 4.16, pp. 879–891. issn: 1742-5689. doi: 10.1098/rsif.2007.1100. url: https://www.ncbi.nlm.nih.gov/pmc/articles/PMC2394553/ (visited on 04/01/2020).

Bansal, Shweta, Jonathan Read, et al. (Sept. 2010). “The dynamic nature of contact networks in infectious disease epidemiology”. In: Journal of Biological Dynamics 4.5. Publisher: Taylor & Francis eprint: 10.1080/17513758.2010.503376, pp. 478–489. issn: 1751-3758. doi: 10.1080/17513758.2010.503376. url: 10.1080/17513758.2010.503376 (visited on 08/01/2023).

Bolker, BM and BT Grenfell (Jan. 1993). “Chaos and biological complexity in measles dynamics”. en. In: Proceedings of the Royal Society of London. Series B: Biological Sciences 251.1330, pp. 75–81. issn: 0962-8452, 1471-2954. doi: 10.1098/rspb.1993.0011. url: https://royalsocietypublishing.org/doi/10.1098/rspb.1993.0011 (visited on 06/05/2023).

Bonabeau, Eric (May 2002). “Agent-based modeling: Methods and techniques for simulating human systems”. en. In: Proceedings of the National Academy of Sciences 99.suppl 3. Publisher: National Academy of Sciences Section: Colloquium Paper, pp. 7280–7287. issn: 0027-8424, 1091-6490. doi: 10.1073/pnas.082080899. url: https://www.pnas.org/content/99/suppl_3/7280 (visited on 09/01/2021).

Bruch, Elizabeth and Jon Atwell (May 2015). “Agent-Based Models in Empirical Social Research”. In: Sociological Methods & Research 44.2. Publisher: SAGE Publications Inc, pp. 186–221. issn: 0049-1241. doi: 10.1177/0049124113506405. url: 10.1177/0049124113506405 (visited on 09/01/2021).

Buckee, Caroline O., Andrew J. Tatem, and C. Jessica E. Metcalf (Jan. 2017). “Seasonal Population Movements and the Surveillance and Control of Infectious Diseases”. en. In: Trends in Parasitology 33.1, pp. 10–20. issn: 1471-4922. doi: 10.1016/j.pt.2016.10.006. url: https://www.sciencedirect.com/science/article/pii/S1471492216301891 (visited on 06/05/2023).

Buitrago-Garcia, Diana et al. (May 2022). “Occurrence and transmission potential of asymptomatic and presymptomatic SARS-CoV-2 infections: Update of a living systematic review and meta-analysis”. en. In: PLOS Medicine 19.5. Publisher: Public Library of Science, e1003987. issn: 1549–1676. doi: 10.1371/journal.pmed.1003987. url: https://journals.plos.org/plosmedicine/article?id=10.1371/journal.pmed.1003987 (visited on 07/04/2023).

Buonomo, B., N. Chitnis, and A. d’Onofrio (June 2018). “Seasonality in epidemic models: a literature review”. en. In: *Ricerche di Matematica* 67.1, pp. 7–25. issn: 1827-3491. doi: 10.1007/s11587-017-0348-6. url: 10.1007/s11587-017-0348-6 (visited on 05/22/2023).

Centers for Disease Control and Prevention (Oct. 2021). Science Brief: SARS-CoV-2 Infection-induced and Vaccine-induced Immunity. en-us. url: https://www.cdc.gov/coronavirus/2019-ncov/science/science-briefs/vaccine-induced-immunity.html (visited on 07/04/2023).

Chin, Alex W H et al. (May 2020). “Stability of SARS-CoV-2 in different environmental conditions”. en. In: The Lancet Microbe 1.1, e10. issn: 2666-5247. doi: 10.1016/S2666-5247(20)30003-3. url: https://www.sciencedirect.com/science/article/pii/S2666524720300033 (visited on 06/05/2023).

Clarke, Kristie E. N. (2022). “Seroprevalence of Infection-Induced SARS-CoV-2 Antibodies — United States, September 2021–February 2022”. en-us. In: *MMWR*. Morbidity and Mortality Weekly Report 71. issn: 0149-21951545-861X. doi: 10.15585/mmwr.mm7117e3. url: https://www.cdc.gov/mmwr/volumes/71/wr/mm7117e3.htm (visited on 02/27/2023).

Crane, Matthew A., et al. (Mar. 2021). “Change in Reported Adherence to Nonpharmaceutical Interventions During the COVID-19 Pandemic, April-November 2020”. In: JAMA 325.9, pp. 883–885. issn: 0098-7484. doi: 10.1001/jama.2021.0286. url: 10.1001/jama.2021.0286 (visited on 02/27/2023).

Csárdi, G. and Nepusz, T. (2006). “The igraph software package for complex network research”. In: Inter-Journal Complex Systems, p. 1695. url: https://cir.nii.ac.jp/crid/1370294643763138711 (visited on 07/06/2023).

Csárdi, Gábor, et al. (June 2023). *igraph*. doi: 10.5281/ZENODO.3630268. url: https://zenodo.org/record/3630268 (visited on 07/06/2023).

Cuevas, Erik (June 2020). “An agent-based model to evaluate the COVID-19 transmission risks in facilities”. en. In: Computers in Biology and Medicine 121, p. 103827. issn: 0010-4825. doi: 10.1016/j.compbiomed.2020.103827. url: https://www.sciencedirect.com/science/article/pii/S001048252030192X (visited on 11/16/2022).

Dabisch, Paul, et al. (Feb. 2021). “The influence of temperature, humidity, and simulated sunlight on the infectivity of SARS-CoV-2 in aerosols”. In: Aerosol Science and Technology 55.2. Publisher: Taylor & Francis eprint: 10.1080/02786826.2020.1829536, pp. 142–153. issn: 0278-6826. doi: 10.1080/02786826.2020.1829536. url: 10.1080/02786826.2020.1829536 (visited on 06/05/2023).

Damette, Olivier, Cĺement Mathonnat, and Stéphane Goutte (June 2021). “Meteorological factors against COVID-19 and the role of human mobility”. en. In: PLOS ONE 16.6. Publisher: Public Library of Science, e0252405. issn: 1932-6203. doi: 10.1371/journal.pone.0252405. url: https://journals.plos.org/plosone/article?id=10.1371/journal.pone.0252405 (visited on 06/05/2023).

Danon, Leon, et al. (Mar. 2011). “Networks and the Epidemiology of Infectious Disease”. en. In: Interdisciplinary Perspectives on Infectious Diseases 2011. Publisher: Hindawi, e284909. issn: 1687-708X. doi: 10.1155/2011/284909. url:https://www.hindawi.com/journals/ipid/2011/284909/ (visited on 09/01/2021).

Davies, Nicholas G., et al. (Apr. 2021). “Estimated transmissibility and impact of SARS-CoV-2 lineage B.1.1.7 in England”. en. In: Science 372.6538. Publisher: American Association for the Advancement of Science Section: Research Article. issn: 0036-8075, 1095-9203. doi: 10.1126/science.abg3055. url: https://science.sciencemag.org/content/372/6538/eabg3055 (visited on 05/05/2021).

Dawson, Daniel E., et al. (Oct. 2018). “Investigating behavioral drivers of seasonal Shiga-Toxigenic Escherichia Coli (STEC) patterns in grazing cattle using an agent-based model”. en. In: PLOS ONE 13.10. Publisher: Public Library of Science, e0205418. issn: 1932-6203. doi: 10.1371/journal.pone.0205418. url: https://journals.plos.org/plosone/article?id=10.1371/journal.pone.0205418 (visited on 06/05/2023).

Dowell, S F (2001). “Seasonal variation in host susceptibility and cycles of certain infectious diseases.” In: Emerging Infectious Diseases 7.3, pp. 369–374. issn: 1080-6040. url: https://www.ncbi.nlm.nih.gov/pmc/articles/PMC2631809/ (visited on 05/22/2023).

Feehan, Dennis M. and Ayesha S. Mahmud (Feb. 2021). “Quantifying population contact patterns in the United States during the COVID-19 pandemic”. en. In: Nature Communications 12.1. Number: 1 Publisher: Nature Publishing Group, p. 893. issn: 2041-1723. doi: 10.1038/s41467-021-20990-2. url: https://www.nature.com/articles/s41467-021-20990-2 (visited on 05/06/2021).

Finkenstädt, B. F. and B. T. Grenfell (2000). “Time series modelling of childhood diseases: a dynamical systems approach”. en. In: Journal of the Royal Statistical Society: Series C (Applied Statistics) 49.2. eprint: https://rss.onlinelibrary.wiley.com/doi/pdf/10.1111/1467-9876.00187, pp. 187–205. issn: 1467-9876. doi: 10.1111/1467-9876.00187. url: https://rss.onlinelibrary.wiley.com/doi/abs/10.1111/1467-9876.00187 (visited on 06/15/2020).

Fisman, D. (Oct. 2012). “Seasonality of viral infections: mechanisms and unknowns”. en. In: Clinical Microbiology and Infection 18.10, pp. 946–954. issn: 1198-743X. doi: 10.1111/j.1469-0691.2012.03968.x. url: https://www.sciencedirect.com/science/article/pii/S1198743X14610910 (visited on 06/28/2023).

Gomez, Jonatan, et al. (Feb. 2021). “INFEKTA—An agent-based model for transmission of infectious diseases: The COVID-19 case in Bogotá, Colombia”. en. In: PLOS ONE 16.2. Publisher: Public Library of Science, e0245787. issn: 1932–6203. doi: 10.1371/journal.pone.0245787. url: https://journals.plos.org/plosone/article?id=10.1371/journal.pone.0245787 (visited on 09/01/2021).

Grassly, Nicholas C and Christophe Fraser (Oct. 2006). “Seasonal infectious disease epidemiology”. In: Proceedings of the Royal Society B: Biological Sciences 273.1600, pp. 2541–2550. issn: 0962-8452. doi: 10.1098/rspb.2006.3604. url: https://www.ncbi.nlm.nih.gov/pmc/articles/PMC1634916/ (visited on 06/15/2020).

He, Daihai, Edward L. Ionides, and Aaron A. King (Feb. 2010). “Plug-and-play inference for disease dynamics: measles in large and small populations as a case study”. In: Journal of the Royal Society Interface 7.43, pp. 271–283. issn: 1742–5689. doi: 10.1098/rsif.2009.0151. url: https://www.ncbi.nlm.nih.gov/pmc/articles/PMC2842609/ (visited on 08/16/2020).

Held, Leonhard and Michaela Paul (2012). “Modeling seasonality in space-time infectious disease surveillance data”. en. In: Biometrical Journal 54.6. eprint: https://onlinelibrary.wiley.com/doi/pdf/10.1002/bimj.201200037, pp. 824–843. issn: 1521-4036. doi: 10.1002/bimj.201200037. url: https://onlinelibrary.wiley.com/doi/abs/10.1002/bimj.201200037 (visited on 05/23/2023).

Hinch, Robert et al. (July 2021). “OpenABM-Covid19—An agent-based model for non-pharmaceutical interventions against COVID-19 including contact tracing”. en. In: PLOS Computational Biology 17.7. Publisher: Public Library of Science, e1009146. issn: 1553-7358. doi: 10.1371/journal.pcbi.1009146. url: https://journals.plos.org/ploscompbiol/article?id=10.1371/journal.pcbi.1009146 (visited on 09/01/2021).

Hoertel, Nicolas, et al. (Sept. 2020). “A stochastic agent-based model of the SARS-CoV-2 epidemic in France”. en. In: *Nature Medicine* 26.9. Bandiera abtest: a Cg type: Nature Research Journals Number: 9 Primary atype: Research Publisher: Nature Publishing Group Subject term: Diseases;Health care Subject term id: diseases;health-care, pp. 1417–1421. issn: 1546-170X. doi: 10.1038/s41591-020-1001-6. url: https://www.nature.com/articles/s41591-020-1001-6 (visited on 09/01/2021).

Holmdahl, Inga, et al. (Nov. 2020). *Frequent testing and immunity-based staffing will help mitigate outbreaks in nursing home settings*. en. Tech. rep. Type: article. medRxiv, p. 2020.11.04.20224758. doi: 10.1101/2020.11.04.20224758. url: https://www.medrxiv.org/content/10.1101/2020.11.04.20224758v2 (visited on 03/04/2022).

Hunter, Elizabeth and John D. Kelleher (June 2021). “Adapting an Agent-Based Model of Infectious Disease Spread in an Irish County to COVID-19”. en. In: Systems 9.2, p. 41. issn: 2079-8954. doi: 10.3390/systems9020041. url: https://www.mdpi.com/2079-8954/9/2/41 (visited on 09/01/2021).

Hunter, Elizabeth, Brian Mac Namee, and John D. Kelleher (2017). “A Taxonomy for Agent-Based Models in Human Infectious Disease Epidemiology”. In: Journal of Artificial Societies and Social Simulation 20.3, p. 2. issn: 1460-7425.

Hunter, Elizabeth, Brian Mac Namee, and John Kelleher (Dec. 2018). “An open-data-driven agent-based model to simulate infectious disease outbreaks”. en. In: PLOS ONE 13.12. Publisher: Public Library of Science, e0208775. issn: 1932-6203. doi: 10.1371/journal.pone.0208775. url: https://journals.plos.org/plosone/article?id=10.1371/journal.pone.0208775 (visited on 09/01/2021).

Johansson, Michael A., et al. (Jan. 2021). “SARS-CoV-2 Transmission From People Without COVID-19 Symptoms”. In: JAMA Network Open 4.1, e2035057. issn: 2574-3805. doi: 10.1001/jamanetworkopen.2020.35057. url: 10.1001/jamanetworkopen.2020.35057 (visited on 11/28/2022).

Keeling, Matt J and Ken T.D Eames (Sept. 2005). “Networks and epidemic models”. In: Journal of The Royal Society Interface 2.4. Publisher: Royal Society, pp. 295–307. doi: 10.1098/rsif.2005.0051. url: https://royalsocietypublishing.org/doi/10.1098/rsif.2005.0051 (visited on 09/01/2021).

Keeling, Matt J., Pejman Rohani, and Bryan T. Grenfell (Jan. 2001). “Seasonally forced disease dynamics explored as switching between attractors”. en. In: Physica D: Nonlinear Phenomena 148.3–4, pp. 317–335. issn: 01672789. doi: 10.1016/S0167-2789(00)00187-1. url: https://linkinghub.elsevier.com/retrieve/pii/S0167278900001871 (visited on 06/05/2023).

Kerr, Cliff C., et al. (Apr. 2021). “Covasim: an agent-based model of COVID-19 dynamics and interventions”. en. In: Company: Cold Spring Harbor Laboratory Press Distributor: Cold Spring Harbor Laboratory Press Label: Cold Spring Harbor Laboratory Press Type: article, p. 2020.05.10.20097469. doi: 10.1101/2020.05.10.20097469. url: https://www.medrxiv.org/content/10.1101/2020.05.10.20097469v3 (visited on 09/01/2021).

Krivorotko, Olga, et al. (Mar. 2022). “Agent-based modeling of COVID-19 outbreaks for New York state and UK: Parameter identification algorithm”. en. In: Infectious Disease Modelling 7.1, pp. 30–44. issn: 2468-0427. doi: 10.1016/j.idm.2021.11.004. url: https://www.sciencedirect.com/science/article/pii/S2468042721000798 (visited on 06/05/2023).

Kronfeld-Schor, N., et al. (Feb. 2021). “Drivers of Infectious Disease Seasonality: Potential Implications for COVID-19”. en. In: Journal of Biological Rhythms 36.1. Publisher: SAGE Publications Inc, pp. 35–54. issn: 0748-7304. doi: 10.1177/0748730420987322. url: 10.1177/0748730420987322 (visited on 05/22/2023).

Levin, Einav G., et al. (Dec. 2021). “Waning Immune Humoral Response to BNT162b2 Covid-19 Vaccine over 6 Months”. In: New England Journal of Medicine 385.24. Publisher: Massachusetts Medical Society eprint: 10.1056/NEJMoa2114583, e84. issn: 0028-4793. doi: 10.1056/NEJMoa2114583. url: 10.1056/NEJMoa2114583 (visited on 07/06/2023).

Liu, Jiangtao et al. (July 2020). “Impact of meteorological factors on the COVID-19 transmission: A multicity study in China”. en. In: Science of The Total Environment 726, p. 138513. issn: 0048-9697. doi: 10.1016/j.scitotenv.2020.138513. url: https://www.sciencedirect.com/science/article/pii/S004896972032026X (visited on 06/05/2023).

Liu, Xiaoyue, et al. (Apr. 2021). “The role of seasonality in the spread of COVID-19 pandemic”. In: Environmental Research 195, p. 110874. issn: 0013-9351. doi: 10.1016/j.envres.2021.110874. url: https://www.ncbi.nlm.nih.gov/pmc/articles/PMC7892320/ (visited on 03/03/2023).

Lofgren, Eric et al. (June 2007). “Influenza Seasonality: Underlying Causes and Modeling Theories”. In: Journal of Virology 81.11. Publisher: American Society for Microbiology, pp. 5429–5436. doi: 10.1128/JVI.01680-06. url: https://journals.asm.org/doi/10.1128/JVI.01680-06 (visited on 02/01/2023).

Lowen, Anice C. and John Steel (July 2014). “Roles of Humidity and Temperature in Shaping Influenza Seasonality”. In: Journal of Virology 88.14, pp. 7692–7695. issn: 0022-538X. doi: 10.1128/JVI.03544-13. url: https://www.ncbi.nlm.nih.gov/pmc/articles/PMC4097773/ (visited on 02/01/2023).

Ma, Yiqun et al. (June 2021). “Role of meteorological factors in the transmission of SARS-CoV-2 in the United States”. en. In: Nature Communications 12.1. Number: 1 Publisher: Nature Publishing Group, p. 3602. issn: 2041-1723. doi: 10.1038/s41467-021-23866-7. url: https://www.nature.com/articles/s41467-021-23866-7 (visited on 06/05/2023).

Marr, Linsey C., et al. (Jan. 2019). “Mechanistic insights into the effect of humidity on airborne influenza virus survival, transmission and incidence”. In: Journal of The Royal Society Interface 16.150. Publisher: Royal Society, p. 20180298. doi: 10.1098/rsif.2018.0298. url: https://royalsocietypublishing.org/doi/10.1098/rsif.2018.0298 (visited on 06/05/2023).

Martin, Joyce, Brady Hamilton, and Michelle Osterman (Aug. 2022). Births in the United States, 2021. Tech. rep. National Center for Health Statistics (U.S.) doi: 10.15620/cdc:119632. url: https://stacks.cdc.gov/view/cdc/119632 (visited on 07/01/2023).

Matson, M. Jeremiah, et al. (2020). “Effect of Environmental Conditions on SARS-CoV-2 Stability in Human Nasal Mucus and Sputum - Volume 26, Number 9—September 2020 - Emerging Infectious Diseases journal - CDC”. en-us. In: doi: 10.3201/eid2609.202267. url: https://wwwnc.cdc.gov/eid/article/26/9/20-2267_article (visited on 06/05/2023).

Mecenas, Paulo, et al. (Sept. 2020). “Effects of temperature and humidity on the spread of COVID-19: A systematic review”. en. In: PLOS ONE 15.9. Publisher: Public Library of Science, e0238339. issn: 1932-6203. doi: 10.1371/journal.pone.0238339. url: https://journals.plos.org/plosone/article?id=10.1371/journal.pone.0238339 (visited on 06/05/2023).

Metcalf, C. Jessica E., et al. (Dec. 2009). “Seasonality and comparative dynamics of six childhood infections in pre-vaccination Copenhagen”. In: Proceedings of the Royal Society B: Biological Sciences 276.1676, pp. 4111–4118. issn: 0962-8452. doi: 10.1098/rspb.2009.1058. url: https://www.ncbi.nlm.nih.gov/pmc/articles/PMC2821338/ (visited on 06/15/2020).

Mistry, D et al. (2021). *SynthPops: a generative model of human contact networks.* original-date: 2020-03-22T19:40:51Z. url: https://github.com/InstituteforDiseaseModeling/synthpops (visited on 11/30/2022).

Moghadas, Seyed M, et al. (Dec. 2021). “The Impact of Vaccination on Coronavirus Disease 2019 (COVID-19) Outbreaks in the United States”. In: Clinical Infectious Diseases 73.12, pp. 2257–2264. issn: 1058-4838. doi: 10.1093/cid/ciab079. url: 10.1093/cid/ciab079 (visited on 11/16/2022).

Morris, Dylan H, et al. (Apr. 2021). “Mechanistic theory predicts the effects of temperature and humidity on inactivation of SARS-CoV-2 and other enveloped viruses”. In: eLife 10. Ed. by Wendy S Garrett, C Brandon Ogbunugafor, and Andreas Handel. Publisher: eLife Sciences Publications, Ltd, e65902. issn: 2050-084X. doi: 10.7554/eLife.65902. url: 10.7554/eLife.65902 (visited on 06/05/2023).

Mossong, Jöel, et al. (Mar. 2008). “Social Contacts and Mixing Patterns Relevant to the Spread of Infectious Diseases”. en. In: PLOS Medicine 5.3. Publisher: Public Library of Science, e74. issn: 1549-1676. doi: 10.1371/journal.pmed.0050074. url: https://journals.plos.org/plosmedicine/article?id=10.1371/journal.pmed.0050074 (visited on 04/01/2020).

National Center for Immunization and Respiratory Diseases (NCIRD) (2022a). National Immunization Survey Adult COVID Module (NIS-ACM). url: https://data.cdc.gov/Vaccinations/National-Immunization-Survey-Adult-COVID-Module-NI/udsf-9v7b (visited on 02/02/2022).

National Center for Immunization and Respiratory Diseases (NCIRD) (2022b). National Immunization Survey Child COVID Module (NIS-CCM). url: https://data.cdc.gov/Vaccinations/National-Immunization-Survey-Child-COVID-Module-NI/uny6-e3dx (visited on 02/02/2022).

National Institutes of Health (2022). COVID-19 Treatment Guidelines Panel: Clinical Spectrum of SARS-CoV-2 Infection. Coronavirus Disease 2019 (COVID-19) Treatment Guidelines. en. url: https://www.covid19treatmentguidelines.nih.gov/overview/clinical-spectrum/ (visited on 02/27/2023).

Nichols, G. L., et al. (Oct. 2021). “Coronavirus seasonality, respiratory infections and weather”. In: BMC Infectious Diseases 21.1, p. 1101. issn: 1471-2334. doi: 10.1186/s12879-021-06785-2. url: 10.1186/s12879-021-06785-2 (visited on 02/01/2023).

Nottmeyer, Luise N. and Francesco Sera (May 2021). “Influence of temperature, and of relative and absolute humidity on COVID-19 incidence in England - A multi-city time-series study”. en. In: Environmental Research 196, p. 110977. issn: 0013-9351. doi: 10.1016/j.envres.2021.110977. url: https://www.sciencedirect.com/science/article/pii/S0013935121002711 (visited on 06/05/2023).

Oraby, Tamer, et al. (Jan. 2014). “Modeling seasonal behavior changes and disease transmission with application to chronic wasting disease”. en. In: Journal of Theoretical Biology 340, pp. 50–59. issn: 0022-5193. doi: 10.1016/j.jtbi.2013.09.003. url: https://www.sciencedirect.com/science/article/pii/S0022519313004244 (visited on 06/05/2023).

Raiteux, Jérémy, et al. (July 2021). “Inactivation of SARS-CoV-2 by Simulated Sunlight on Contaminated Surfaces”. In: Microbiology Spectrum 9.1. Publisher: American Society for Microbiology, 10.1128/spectrum.00333–21. doi: 10.1128/spectrum.00333-21. url: https://journals.asm.org/doi/10.1128/Spectrum.00333-21 (visited on 06/05/2023).

Reicher, Stephen and John Drury (Jan. 2021). “Pandemic fatigue? How adherence to covid-19 regulations has been misrepresented and why it matters”. en. In: BMJ 372. Publisher: British Medical Journal Publishing Group Section: Views &amp; Reviews, n137. issn: 1756-1833. doi: 10.1136/bmj.n137. url: https://www.bmj.com/content/372/bmj.n137 (visited on 02/27/2023).

Riddell, Shane, et al. (Oct. 2020). “The effect of temperature on persistence of SARS-CoV-2 on common surfaces”. In: Virology Journal 17.1, p. 145. issn: 1743-422X. doi: 10.1186/s12985-020-01418-7. url: 10.1186/s12985-020-01418-7 (visited on 06/05/2023).

Roubenoff, Ethan, Dennis M. Feehan, and Ayesha S. Mahmud (June 2023). “Evaluating primary and booster vaccination prioritization strategies for COVID-19 by age and high-contact employment status using data from contact surveys”. en. In: Epidemics 43, p. 100686. issn: 1755-4365. doi: 10.1016/j.epidem.2023.100686. url: https://www.sciencedirect.com/science/article/pii/S1755436523000221 (visited on 06/27/2023).

Sah, Pratha et al. (May 2021). “Accelerated vaccine rollout is imperative to mitigate highly transmissible COVID-19 variants”. eng. In: EClinicalMedicine 35, p. 100865. issn: 2589–5370. doi: 10.1016/j.eclinm.2021.100865.

Sera, Francesco, et al. (Oct. 2021). “A cross-sectional analysis of meteorological factors and SARS-CoV-2 transmission in 409 cities across 26 countries”. en. In: Nature Communications 12.1. Number: 1 Publisher: Nature Publishing Group, p. 5968. issn: 2041-1723. doi: 10.1038/s41467-021-25914-8. url: https://www.nature.com/articles/s41467-021-25914-8 (visited on 06/05/2023).

Shaman, Jeffrey and Melvin Kohn (Mar. 2009). “Absolute humidity modulates influenza survival, transmission, and seasonality”. In: Proceedings of the National Academy of Sciences 106.9. Publisher: Proceedings of the National Academy of Sciences, pp. 3243–3248. doi: 10.1073/pnas.0806852106. url: https://www.pnas.org/doi/full/10.1073/pnas.0806852106 (visited on 06/05/2023).

Silk, Benjamin J. (2023). “COVID-19 Surveillance After Expiration of the Public Health Emergency Declaration — United States, May 11, 2023”. en-us. In: *MMWR*. Morbidity and Mortality Weekly Report 72. issn: 0149-21951545-861X. doi: 10.15585/mmwr.mm7219e1. url: https://www.cdc.gov/mmwr/volumes/72/wr/mm7219e1.htm (visited on 06/30/2023).

Smith, Thomas P. et al. (June 2021). “Temperature and population density influence SARS-CoV-2 transmission in the absence of nonpharmaceutical interventions”. In: Proceedings of the National Academy of Sciences 118.25. Publisher: Proceedings of the National Academy of Sciences, e2019284118. doi: 10.1073/pnas.2019284118. url: https://www.pnas.org/doi/full/10.1073/pnas.2019284118 (visited on 06/05/2023).

Strasser, Zachary H., et al. (Oct. 2022). “Estimates of SARS-CoV-2 Omicron BA.2 Subvariant Severity in New England”. In: JAMA Network Open 5.10, e2238354. issn: 2574-3805. doi: 10.1001/jamanetworkopen.2022.38354. url: 10.1001/jamanetworkopen.2022.38354 (visited on 02/27/2023).

Susswein, Zachary, Eva C Rest, and Shweta Bansal (Apr. 2023). “Disentangling the rhythms of human activity in the built environment for airborne transmission risk: An analysis of large-scale mobility data”. In: eLife 12. Ed. by Niel Hens, Diane M Harper, and Guillaume Béraud. Publisher: eLife Sciences Publications, Ltd, e80466. issn: 2050-084X. doi: 10.7554/eLife.80466. url: 10.7554/eLife.80466 (visited on 08/01/2023).

Tenforde, Mark W. (2021). “Effectiveness of Pfizer-BioNTech and Moderna Vaccines Against COVID-19 Among Hospitalized Adults Aged *≥*65 Years — United States, January–March 2021”. en-us. In: *MMWR*. Morbidity and Mortality Weekly Report 70. issn: 0149-21951545-861X. doi: 10.15585/mmwr.mm7018e1. url: https://www.cdc.gov/mmwr/volumes/70/wr/mm7018e1.htm (visited on 09/07/2021).

Thompson, Mark G. (2021). “Interim Estimates of Vaccine Effectiveness of BNT162b2 and mRNA-1273 COVID-19 Vaccines in Preventing SARS-CoV-2 Infection Among Health Care Personnel, First Responders, and Other Essential and Frontline Workers — Eight U.S. Locations, December 2020–March 2021”. en-us. In: *MMWR*. Morbidity and Mortality Weekly Report 70. issn: 0149-21951545-861X. doi: 10.15585/mmwr.mm7013e3. url: https://www.cdc.gov/mmwr/volumes/70/wr/mm7013e3.htm (visited on 09/07/2021).

Thompson, Mark G. (2022). “Effectiveness of a Third Dose of mRNA Vaccines Against COVID-19–Associated Emergency Department and Urgent Care Encounters and Hospitalizations Among Adults During Periods of Delta and Omicron Variant Predominance — VISION Network, 10 States, August 2021–January 2022”. en-us. In: *MMWR*. Morbidity and Mortality Weekly Report 71. issn: 0149-21951545-861X. doi: 10.15585/mmwr.mm7104e3. url: https://www.cdc.gov/mmwr/volumes/71/wr/mm7104e3.htm (visited on 03/02/2022).

US Census Bureau (2022). Table B01001: Sex by Age, 2021. American Community Survey 1-year Estimates. url: https://data.census.gov/table?q=B01001:+SEX+BY+AGE&tid=ACSDT1Y2021.B01001 (visited on 07/06/2023).

Venkatramanan, Srinivasan, et al. (Mar. 2018). “Using data-driven agent-based models for forecasting emerging infectious diseases”. en. In: Epidemics. The RAPIDD Ebola Forecasting Challenge 22, pp. 43–49. issn: 1755-4365. doi: 10.1016/j.epidem.2017.02.010. url: https://www.sciencedirect.com/science/article/pii/S1755436517300221 (visited on 09/01/2021).

Weaver, Amanda K. et al. (2022). “Environmental Factors Influencing COVID-19 Incidence and Severity”. In: Annual Review of Public Health 43.1. eprint: 10.1146/annurev-publhealth-052120-101420, pp. 271–291. doi: 10.1146/annurev-publhealth-052120-101420. url: 10.1146/annurev-publhealth-052120-101420 (visited on 06/05/2023).

Williams, Katherine V., et al. (Nov. 2022). “Can a Two-Dose Influenza Vaccine Regimen Better Protect Older Adults? An Agent-Based Modeling Study”. en. In: Vaccines 10.11. Number: 11 Publisher: Multidisciplinary Digital Publishing Institute, p. 1799. issn: 2076-393X. doi: 10.3390/vaccines10111799. url: https://www.mdpi.com/2076-393X/10/11/1799 (visited on 06/05/2023).

Wu, Yu, et al. (Aug. 2020). “Effects of temperature and humidity on the daily new cases and new deaths of COVID-19 in 166 countries”. en. In: Science of The Total Environment 729, p. 139051. issn: 0048-9697. doi: 10.1016/j.scitotenv.2020.139051. url: https://www.sciencedirect.com/science/article/pii/S0048969720325687 (visited on 06/05/2023).

Xu, Jiaquan, et al. (Dec. 2022). *Mortality in the United States,* 2021. Tech. rep. National Center for Health Statistics (U.S.) doi: 10.15620/cdc:122516. url: https://stacks.cdc.gov/view/cdc/122516 (visited on 07/01/2023).

Yang, Zhen, et al. (Apr. 2022). “Clinical Characteristics, Transmissibility, Pathogenicity, Susceptible Populations, and Re-infectivity of Prominent COVID-19 Variants”. In: Aging and Disease 13.2, pp. 402–422. issn: 2152-5250. doi: 10.14336/AD.2021.1210. url: https://www.ncbi.nlm.nih.gov/pmc/articles/PMC8947836/ (visited on 02/27/2023).

